# Primary exposure to Zika virus increases risk of symptomatic dengue virus infection with serotypes 2, 3, and 4 but not serotype 1

**DOI:** 10.1101/2023.11.29.23299187

**Authors:** Jose Victor Zambrana, Chloe M. Hasund, Rosemary A. Aogo, Sandra Bos, Sonia Arguello, Karla Gonzalez, Damaris Collado, Tatiana Miranda, Guillermina Kuan, Aubree Gordon, Angel Balmaseda, Leah Katzelnick, Eva Harris

## Abstract

Infection with any of the four dengue virus serotypes (DENV1-4) can protect against or enhance subsequent dengue depending on pre-existing antibodies and the subsequent infecting serotype. Additionally, primary infection with the related flavivirus Zika virus (ZIKV) has been shown to increase DENV2 disease. Here, we measured how prior DENV and ZIKV immunity influenced risk of disease caused by all four serotypes in a pediatric Nicaraguan cohort. Of 3,412 participants in 2022, 10.6% experienced symptomatic DENV infections caused by DENV1 (n=139), DENV4 (n=133), DENV3 (n=54), DENV2 (n=9), or an undetermined serotype (n=39). Longitudinal clinical and serological data were used to define infection histories, and generalized linear and additive models adjusted for age, sex, time since the last infection, cohort year, and repeat measurements were used to predict disease risk. Compared to flavivirus-naïve participants, primary ZIKV infection increased disease risk of DENV4 (relative risk = 2.62, 95% confidence interval: 1.48-4.63) and DENV3 (2.90, 1.34-6.27) but not DENV1 (1.20, 0.72-1.99). Primary DENV infection or a DENV followed by ZIKV infection also increased DENV4 risk. We re-analyzed 19 years of cohort data and demonstrated that prior flavivirus-immunity and pre-existing antibody titer differentially affected disease risk for incoming serotypes, increasing risk of DENV2 and DENV4, protecting against DENV1, and protecting at high titers but enhancing at low titers against DENV3. We thus find that prior ZIKV infection, like prior DENV infection, increases risk of certain DENV serotypes. Cross-reactivity among flaviviruses should be carefully considered when assessing vaccine safety and efficacy.

**One-Sentence Summary:** Dengue disease risk is differentially modulated depending on pre- existing immunity to dengue and Zika virus infections and the secondary infecting serotype.

## INTRODUCTION

Dengue virus, comprised of four distinct serotypes (DENV1-4), and Zika virus (ZIKV) are antigenically related, mosquito-borne flaviviruses that cause a significant global health burden (*1–3*). Both flaviviruses are transmitted by female *Aedes aegypti* mosquitoes, co-circulate in many countries, and cause major epidemics worldwide (*1*). DENV and ZIKV infection each induce antibodies that cross-react with the other viruses, but how these antibodies modulate subsequent disease risk has only been partially elucidated (*4, 5*).

Following a DENV or ZIKV infection, neutralizing antibodies at high titers are observed to provide long-lasting protection against the infecting virus, a phenomenon termed homotypic protection (*6*). Cross-reactive neutralizing antibodies can provide protection against an incoming heterotypic infection (*7–9*). However, DENV infection also elicits low-to-intermediate cross-reactive antibody titers, which can increase risk of a symptomatic infection and enhance disease severity in a subsequent DENV infection with a different serotype (*10–13*). This increased risk has been attributed to a phenomenon known as antibody-dependent enhancement (ADE), whereby non- or poorly neutralizing antibodies facilitate DENV entry into host cells through the Fcγ receptor, improving infection efficiency and activating target immune cells (*14, 15*). Increased risk of future dengue disease severity following a DENV infection is well established (*11, 14, 16*), and two studies have reported an association between prior ZIKV infection and DENV2 disease risk (*17, 18*). This last finding is consistent with studies in macaques exposed to ZIKV and then DENV2, which have shown an increase in viremia compared to ZIKV-naïve macaques (*19, 20*). It is unclear whether primary ZIKV infection modulates secondary dengue caused by other serotypes.

Symptomatic and severe disease occur more frequently in secondary DENV2 and DENV4 infections, as compared to DENV1 and DENV3 infections (*10, 11, 21–24*). In line with this observation, a higher neutralizing antibody titer is needed to protect against symptomatic DENV2 versus other serotypes (*9, 23, 25*). We previously showed that a broad range of pre-existing anti-DENV binding antibody titers can enhance DENV2 disease, low titers can enhance DENV3, and high titers protect against DENV1 and DENV3 (*26*). Less is known about the effect of pre-existing antibody titer on DENV4.

In 2022, all four DENV serotypes co-circulated in Nicaragua in populations affected by the 2016 Zika epidemic. This large dengue epidemic (n=374 cases) in the Nicaraguan Pediatric Cohort Study enabled us to evaluate whether prior ZIKV and DENV infections modulate risk of secondary dengue caused by DENV1, DENV3, or DENV4. We also evaluated whether individuals with a prior DENV infection followed by a ZIKV infection (DENV-ZIKV) had similar outcomes as individuals with a primary ZIKV infection followed by DENV infection (ZIKV-DENV), a group observed for the first time in the Nicaraguan cohort. Further, we measured the effect of flavivirus immunity and pre-existing DENV antibody titers on symptomatic DENV1-4 infection over 19 years of cohort data and performed a mediation analysis to examine the relative contribution of antibody titer to the relationship between infection history and DENV infection outcome. Finally, we evaluated the long-term antibody kinetics in individuals with varied flavivirus infection histories.

## RESULTS

### All four DENV serotypes co-circulated during the 2022-2023 dengue epidemic in Nicaragua, and prior DENV or ZIKV infections increase risk of dengue cases

The Pediatric Dengue Cohort Study has followed ∼4,000 participants ages 2-17 years for arbovirus infection in Managua, Nicaragua, continuously since 2004 (table S1). In 2022, for the first time since the cohort was established, all four DENV serotypes were detected in cases, circulating simultaneously (Fig. 1A). A total of 10.6% of all participants experienced symptomatic DENV infections; 374 cases met the dengue case definition and were confirmed by RT-PCR or paired acute and convalescent serology of the 3,412 active cohort participants with complete infection histories. Most infections were caused by DENV1 (n=139) and DENV4 (n=133), followed by DENV3 (n=54) and DENV2 (n=9), and a subset were confirmed by serology (n=39) (Table 1).

**Fig. 1.**
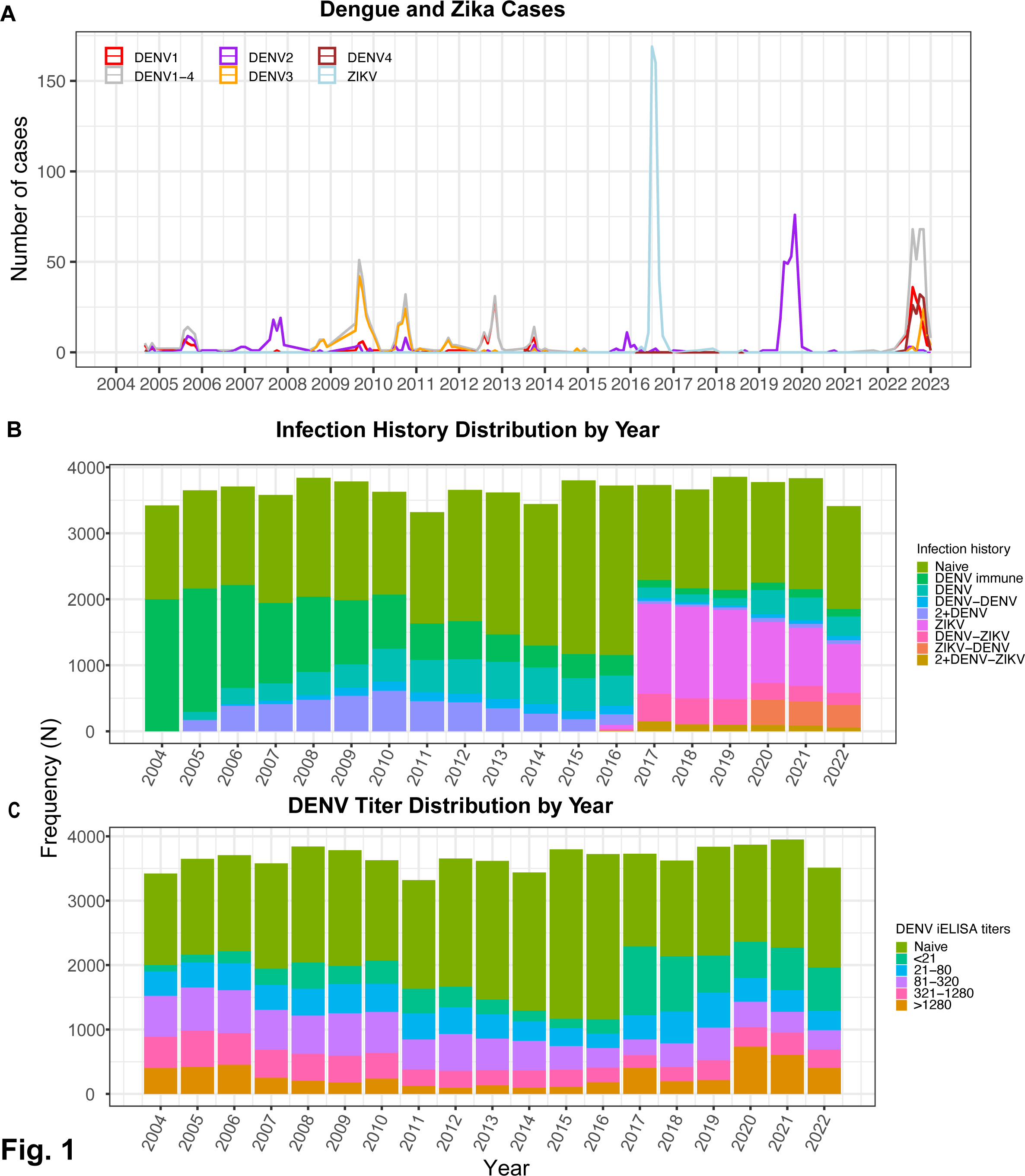
Nicaragua experienced a large dengue epidemic in 2022-2023 with co-circulation of all four serotypes. Monthly dengue and Zika cases **(A)**, the proportion of individuals with various infection histories **(B)**, and DENV iELISA titer distribution **(C)** by epidemic year in the Pediatric Dengue Cohort Study (2004-2022).

**Table. 1.** Characteristics of participants in the Pediatric Dengue Cohort Study with documented infection histories in 2022-23.

| Variable | Total<br>Population | DENV cases |  |  |  |  |  | p-value<sup>2</sup> |
| --- | --- | --- | --- | --- | --- | --- | --- | --- |
|  | N = 3,564<sup>1</sup> | Overall, N<br>= 374<sup>1</sup> | DENV1, N<br>= 139<sup>1</sup> | DENV2,<br>N = 9<sup>1</sup> | DENV3, N<br>= 54<sup>1</sup> | DENV4,<br>N = 133<sup>1</sup> | Dengue<br>serology,<br>N = 39<sup>1</sup> |  |
| Age | 10.4 (4.0) | 11.0 (3.6) | 10.6 (3.8) | 10.2 (4.7) | 11.1 (3.7) | 11.9 (3.3) | 9.7 (3.3) | 0.003 |
| Sex |  |  |  |  |  |  |  | 0.2 |
| F | 1,768 (50%) | 193 (52%) | 64 (46%) | 4 (44%) | 25 (46%) | 78 (59%) | 22 (56%) |  |
| M | 1,794 (50%) | 179 (48%) | 74 (54%) | 5 (56%) | 29 (54%) | 54 (41%) | 17 (44%) |  |
| DHF/DSS | 3 (<0.1%) | 3 (0.8%) | 1 (0.7%) | 0 (0%) | 2 (3.7%) | 0 (0%) | 0 (0%) | 0.2 |
| DwWS/SD | 57 (1.6%) | 57 (15%) | 27 (19%) | 1 (11%) | 4 (7.4%) | 23 (17%) | 2 (5.1%) | 0.076 |
| Time since last<br>infection | 5.50 (3.00) | 5.81<br>(2.87) | 5.90<br>(3.01) | 6.25<br>(3.65) | 6.41<br>(2.97) | 5.35<br>(2.73) | 6.18<br>(2.33) | 0.049 |
| Geometric Mean<br>Titer (iELISA) | 665 (1,985) | 390<br>(1,400) | 314<br>(1,263) | 6 (11) | 58 (198) | 684<br>(1,876) | 154 (563) | <0.001 |
| Infection history |  |  |  |  |  |  |  |  |
| Naive | 1,559 (46%) | 123 (35%) | 55 (43%) | 4 (50%) | 19 (37%) | 24 (19%) | 21 (62%) |  |
| DENV-immune | 113 (3.3%) | 12 (3.4%) | 3 (2.3%) | 0 (0%) | 4 (7.8%) | 4 (3.1%) | 1 (2.9%) |  |
| DENV | 299 (8.8%) | 33 (9.5%) | 11 (8.6%) | 0 (0%) | 1 (2.0%) | 20 (16%) | 1 (2.9%) |  |
| ZIKV | 742 (22%) | 110 (32%) | 37 (29%) | 4 (50%) | 21 (41%) | 41 (32%) | 7 (21%) |  |
| DENV-DENV | 63 (1.8%) | 12 (3.4%) | 7 (5.5%) | 0 (0%) | 1 (2.0%) | 4 (3.1%) | 0 (0%) |  |
| DENV-ZIKV | 174 (5.1%) | 25 (7.2%) | 6 (4.7%) | 0 (0%) | 0 (0%) | 17 (13%) | 2 (5.9%) |  |
| ZIKV-DENV | 334 (9.8%) | 25 (7.2%) | 7 (5.5%) | 0 (0%) | 4 (7.8%) | 13 (10%) | 1 (2.9%) |  |
| 2+DENV | 56 (1.6%) | 2 (0.6%) | 1 (0.8%) | 0 (0%) | 0 (0%) | 1 (0.8%) | 0 (0%) |  |
| 2+DENV-ZIKV | 43 (1.3%) | 6 (1.7%) | 1 (0.8%) | 0 (0%) | 1 (2.0%) | 3 (2.4%) | 1 (2.9%) |  |
| DENV-ZIKV-2+DENV | 3 (<0.1%) | 0 (0%) | 0 (0%) | 0 (0%) | 0 (0%) | 0 (0%) | 0 (0%) |  |
| ZIKV-DENV-<br>DENV | 15 (0.4%) | 0 (0%) | 0 (0%) | 0 (0%) | 0 (0%) | 0 (0%) | 0 (0%) |  |
| 2+DENV-ZIKV-<br>DENV | 11 (0.3%) | 0 (0%) | 0 (0%) | 0 (0%) | 0 (0%) | 0 (0%) | 0 (0%) |  |

<sup>1</sup>Mean (SD); n (%)
<sup>2</sup>Kruskal-Wallis rank sum test; Fisher's exact test

Before the 2022-2023 epidemic in Nicaragua, the ZIKV pandemic affected the cohort in 2016, followed by a large DENV2 epidemic in 2019, with minimal flavivirus circulation observed between these epidemics. Further, DENV2 was the only serotype detected in cases between 2014 and 2021. Thus, based on the young ages of the cohort, nearly half of participants (n=48.6%) were only ever exposed to ZIKV or DENV2. In 2022, 45.7% of the cohort were flavivirus-naïve, 21.7% had experienced only a prior ZIKV infection, 8.8% had only had one prior DENV infection, and 23.8% had experienced different combinations of DENV and ZIKV infections (Fig. 1B). Notably, DENV seropositivity and population anti-DENV antibody titers declined after the DENV3 epidemic in 2010-2011, titers increased after the ZIKV epidemic in 2016, and seropositivity and titers increased again after the DENV2 epidemic in 2019-2020 (Fig. 1B and 1C).

During the 2022-2023 dengue epidemic, the overall risk of experiencing a dengue case was 7.8% (6.7%-9.4%) for the flavivirus-naïve group, our reference group, and was significantly higher for participants with one prior ZIKV infection (risk = 14.8%, 12.5%-17.6%), two prior DENV infections (DENV-DENV) (19.0, 11.4%-31.7%), and one prior DENV and then ZIKV infection (DENV-ZIKV) (14.4%, 10.0%-20.7%) (fig. S1). We were unable to calculate risk for some of the more complex infection histories, including those with different sequences of DENV and ZIKV infections followed by DENV infection (DENV-ZIKV-2+DENV, ZIKV-DENV-DENV, and 2+DENV-ZIKV-DENV), because of low numbers of individuals in these groups and lack of any symptomatic DENV infections.

### Primary DENV and ZIKV infection increases risk of symptomatic infection caused by DENV2 and DENV4 but not DENV1, while primary ZIKV infection also increases risk of symptomatic DENV3 infection

To investigate the role of infection history on outcome by serotype, we first evaluated whether primary ZIKV infection increased the risk of symptomatic infections caused by DENV1, DENV3, and DENV4 during the 2022-2023 epidemic. Only 9 DENV2 cases were detected in 2022, thus we could not analyze DENV2 case risk by infection histories. Using log-binomial generalized linear models (GLMs) adjusted for age, sex, and time since previous infection, we found that primary ZIKV infection significantly increased the risk of symptomatic DENV infection compared to flavivirus-naïve individuals (Risk Ratio (RR) = 1.61, 95% CI: 1.20-2.17). When these analyses were stratified by infecting serotype, risk of disease was increased for DENV3 (RR = 2.90, 1.34-6.27) and DENV4 (RR = 2.62, 1.48-4.63), but not DENV1 (Fig. 2A and table S2). Children with a prior ZIKV infection also trended toward increased risk of dengue with warning signs or severe dengue (DwWS/SD) (RR=2.19, 0.99: 4.83, *p* = 0.053) compared to flavivirus-naïve children.

**Fig. 2.**
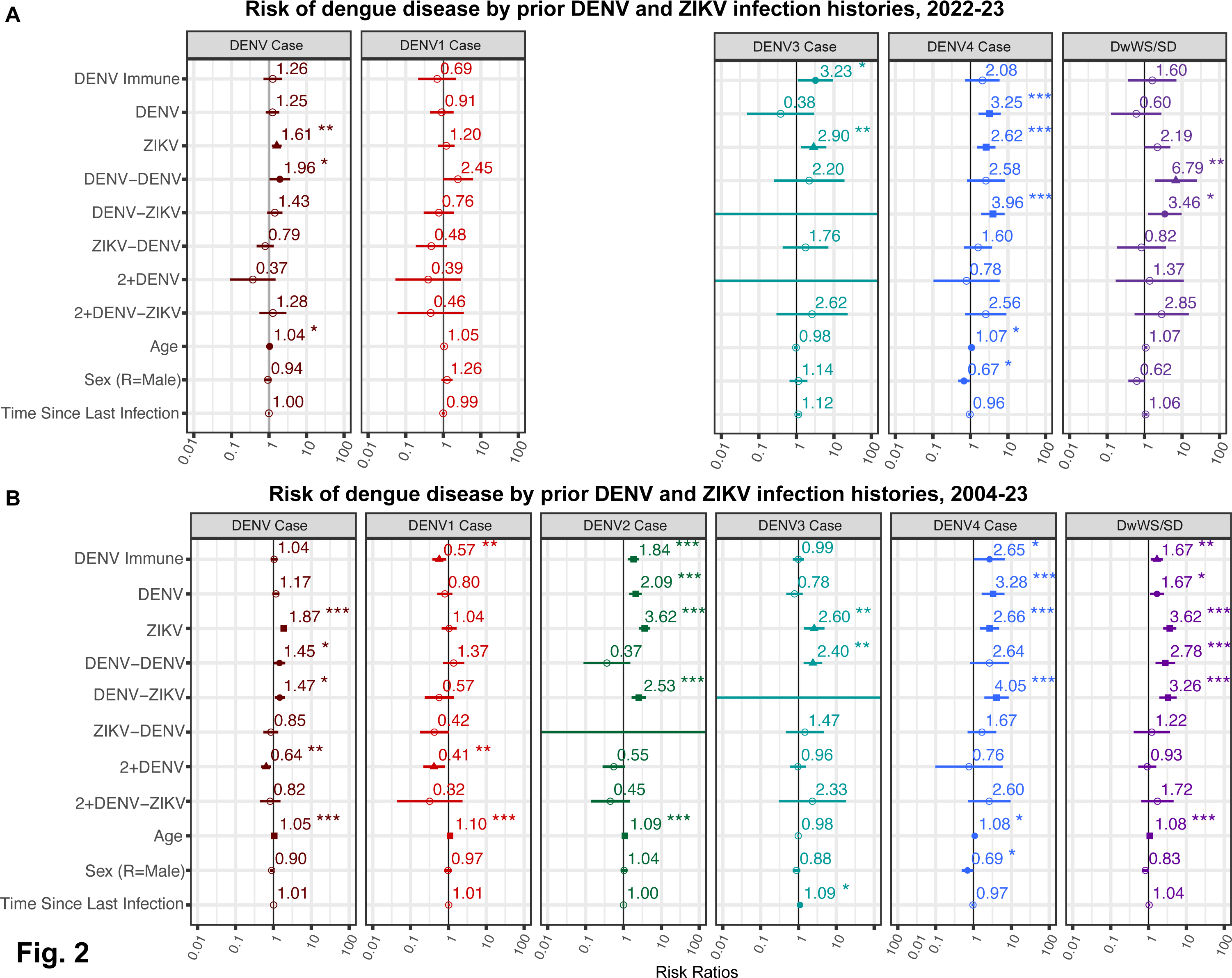
Prior infection history confers differential risk of symptomatic DENV1, DENV3 and DENV4 and overall disease and disease severity. Log-binomial and log-Poisson generalized linear models (GLMs) were used to estimate the risk of any dengue case, a case caused by DENV1, DENV2, DENV3, or DENV4, and Dengue with Warning Signs/Severe Dengue (DwWS/SD) by DENV and ZIKV infection history in **(A)** the 2022-2023 epidemic or **(B)** all years of the cohort study (2004-2023).

To increase statistical power and resolution and to include DENV2, we performed the above analyses on data from the entire Pediatric Dengue Cohort Study timeframe (2004 to 2023). We used both log-binomial and log-poisson generalized linear mixed-effect models (GLMMs) adjusted for age, sex, time since last infection, and cohort epidemic year, with clustering at the individual level to account for repeat measurements. Similar to observations for the 2022-2023 epidemic, we found that compared to flavivirus-naïve individuals, a prior ZIKV infection significantly increased the risk of symptomatic dengue overall (RR=1.87, 1.54-2.27) as well as the risk of DwWS/SD (RR = 3.62, 2.44-5.38) (Fig. 2B and table S3). Primary ZIKV infection significantly increased the risk for a subsequent DENV2, DENV3, and DENV4 infection, but not DENV1.

We performed the same analyses for participants with primary DENV infection. When analyzed across cohort years, primary DENV infection significantly increased the risk of symptomatic DENV2 (RR = 2.09, 1.44-3.05) and DENV4 (RR = 3.28, 1.64-6.57) infections, but not DENV1 or DENV3 infections (Fig. 2B and table S3). Unlike a prior ZIKV infection, a single prior DENV infection did not increase risk of symptomatic dengue overall (RR = 1.17, 0.94-1.45); however, one prior DENV infection did increase overall risk of DwWS/SD (RR = 1.67, 1.09- 2.57), although not as much as prior ZIKV (Fig. 2B and table S3).

### The number and order of DENV and ZIKV infections determine risk of developing future dengue

Sequential infection with different DENV serotypes is thought to protect against disease by inducing broadly protective antibodies against all serotypes (*27*). We previously found that individuals with primary DENV infection followed by a ZIKV infection (DENV-ZIKV) were at increased risk of subsequent symptomatic DENV2 infection compared to being flavivirus-naïve, suggesting that the additional ZIKV infection did not broaden their protective immunity (*26*). Here, we compared dengue disease risk for DENV-ZIKV infection histories to children with a previously unobserved sequence of infections: ZIKV followed by one DENV infection (ZIKV-DENV). In the analysis of all epidemic seasons, the DENV-ZIKV infection history was found to significantly increase the risk of subsequent symptomatic dengue overall (RR = 1.47, 1.09-1.99) and specifically, subsequent symptomatic infection with DENV2 (RR = 2.53, 1.64-3.91) and DENV4 (RR = 4.05, 1.93-8.49), as well as DwWS/SD (RR = 3.26, 1.94-5.50) (Fig. 2B and table S3). In contrast, the ZIKV-DENV infection sequence did not increase the risk of future symptomatic DENV1-4 infections, dengue disease, or DwWS/SD (Fig. 2A and 2B, table S2 and S3).

Counter to our expectation, we also found that across epidemic seasons, those with two prior DENV infections (DENV-DENV) were at increased risk of subsequent symptomatic dengue overall (RR=2.66, 1.49-4.76) and DwWS/SD (3.62, 2.44-5.38) compared to the flavivirus-naïve group (Fig. 2A and 2B, tables S2 and S3). The DENV-DENV group was also at a greater risk of symptomatic DENV3 infection (RR = 2.40, 1.37-4.23) (Fig. 2B and table S3). However, those with two or more DENV infections (2+DENV group) or at least two DENV infections and a ZIKV infection (2+DENV-ZIKV) were not at increased risk of either symptomatic or severe dengue (Fig. 2B and table S3). Protective effects were only observed for the 2+ DENV group against symptomatic DENV infection overall (RR = 0.64, 0.48-0.86), and DENV1 (RR = 0.41, 0.22-0.79). Individuals with multiple flavivirus infections may live in high-risk areas for exposure to DENV. We thus performed a sensitivity analysis where we controlled for the age-adjusted DENV seroprevalence at the neighborhood level in the year prior to the recorded dengue case for each participant and clustered participants by neighborhood. Results were similar to our previous results (Fig. 2) for both the 2022-2023 epidemic season and the complete cohort epidemic seasons (2004–2023) across infection histories, suggesting that spatial risk did not explain the increased risk of disease observed for the DENV-DENV group (fig. S2).

We investigated when the 12 individuals with dengue cases in 2022-2023 in the DENV-DENV group experienced their previous infection to determine the likely prior infecting serotype. Most of those with tertiary DENV4 cases had their second infection three years previously, during the DENV2 epidemic, whereas most DENV1 tertiary cases had experienced their second infection in 2021, right before 2022 epidemic when multiple serotypes may have already been circulating (fig. S3). While some sequential infections detected by DENV iELISA in previous years may have been due to homotypic re-exposure, the epidemiology of dengue in the cohort makes it likely that those experiencing tertiary infections in 2022-2023 were infected with distinct serotypes.

Of note, consistent with previous findings, older age was a significant risk factor for dengue disease as well as for symptomatic DENV1, DENV2 and DENV4 infections in the full cohort analysis. Sex was not a significant factor except with DENV4, where male sex was associated with reduced risk of symptomatic infection in both analyses (Fig. 2A and 2B). Time since infection was not a significant risk factor in any model, except for a slightly increased risk of DENV3 in the complete cohort analysis (Fig. 2A and 2B).

### Pre-existing flavivirus immunity and antibody titers protect against DENV1, but can increase risk of DENV2, DENV3, and DENV4

Across infection histories, we observed increased risk of DENV2, DENV3, and DENV4 disease and DwWS/SD and protective effects against symptomatic DENV1 infection, comparing to flavivirus-naïve individuals. To explore how overall flavivirus immunity affected disease caused by each serotype, we grouped individuals with any prior DENV or ZIKV infection, regardless of serotype or number of infections, as “flavivirus-immune” and the non-exposed as “flavivirus-naïve.” Compared to flavivirus-naïve individuals, those who were flavivirus-immune were at significantly increased risk of dengue disease (*p*=<0.001), DwWS/SD (*p*=<0.001), and DHF/DSS (*p*=<0.001) (Fig. 3 and table S4). However, distinct risks were observed for different serotypes, with significant risk for flavivirus-immune participants of DENV2 (*p*=<0.001) and DENV4 disease (*p*=0.002), but not DENV3 (*p*=0.551). Conversely, flavivirus-immune individuals were at significantly lower risk of DENV1 (*p*=0.036), indicating protection (Fig. 3 and table S5).

**Fig. 3.**
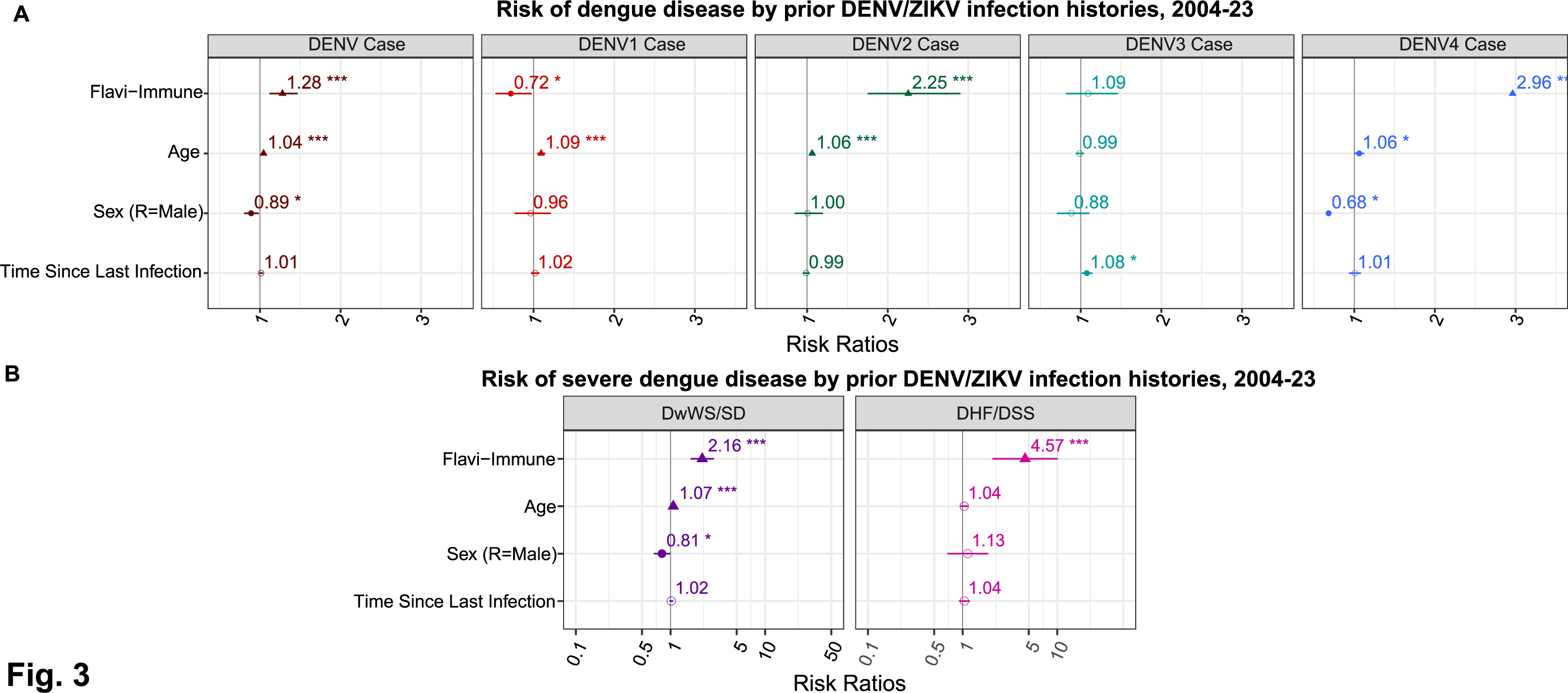
History of prior flavivirus infection exacerbates risk of symptomatic DENV2 and DENV4 and overall disease and disease severity. Linear mixed effects models were used to estimate the risks of a symptomatic DENV infection (case) by any serotype or from and DENV1, DENV2, DENV3, or DENV4 infection **(A)** and Dengue with Warning Signs/Severe Dengue (DwWS/SD) and Dengue Hemorrhagic Fever/Dengue Shock Syndrome (DHF/DSS) **(B)** in the prospective cohort study, 2004-2023 by immune status (naïve or flavivirus immune).

We previously showed that pre-existing antibodies, as measured by iELISA (*12*), can protect or enhance dengue disease and disease severity depending on the titer magnitude and secondary infecting serotype (*12, 26*). With the recent circulation of DENV4 in the region, we were also able to test how pre-existing anti-flavivirus antibodies modulate all four serotypes, both for the 2022 epidemic (Fig. 4A) and across all cohort years (Fig. 4B). Here, we found that DENV iELISA titers (except those >1280) significantly increased risk of symptomatic DENV4 infection compared to flavivirus-naïve children. This effect paralleled that seen for DENV2 in our full cohort analyses. The pattern was different for DENV3, where low titers (<21) increased disease (*p*=0.005) while high titers were protective in the full cohort analyses. Finally, no enhancing effects were observed against DENV1, while higher titers of pre-existing anti-DENV antibodies were protective (*p*=0.041), as seen against DENV3 (Fig. 4A and table S3). Regarding the overall risk of dengue disease and DwWS/SD, we found that low iELISA titers were associated with increased risk of both symptomatic DENV infection and DwWS/SD (Fig. 4 A and B, table S5 and S6).

**Fig. 4.**
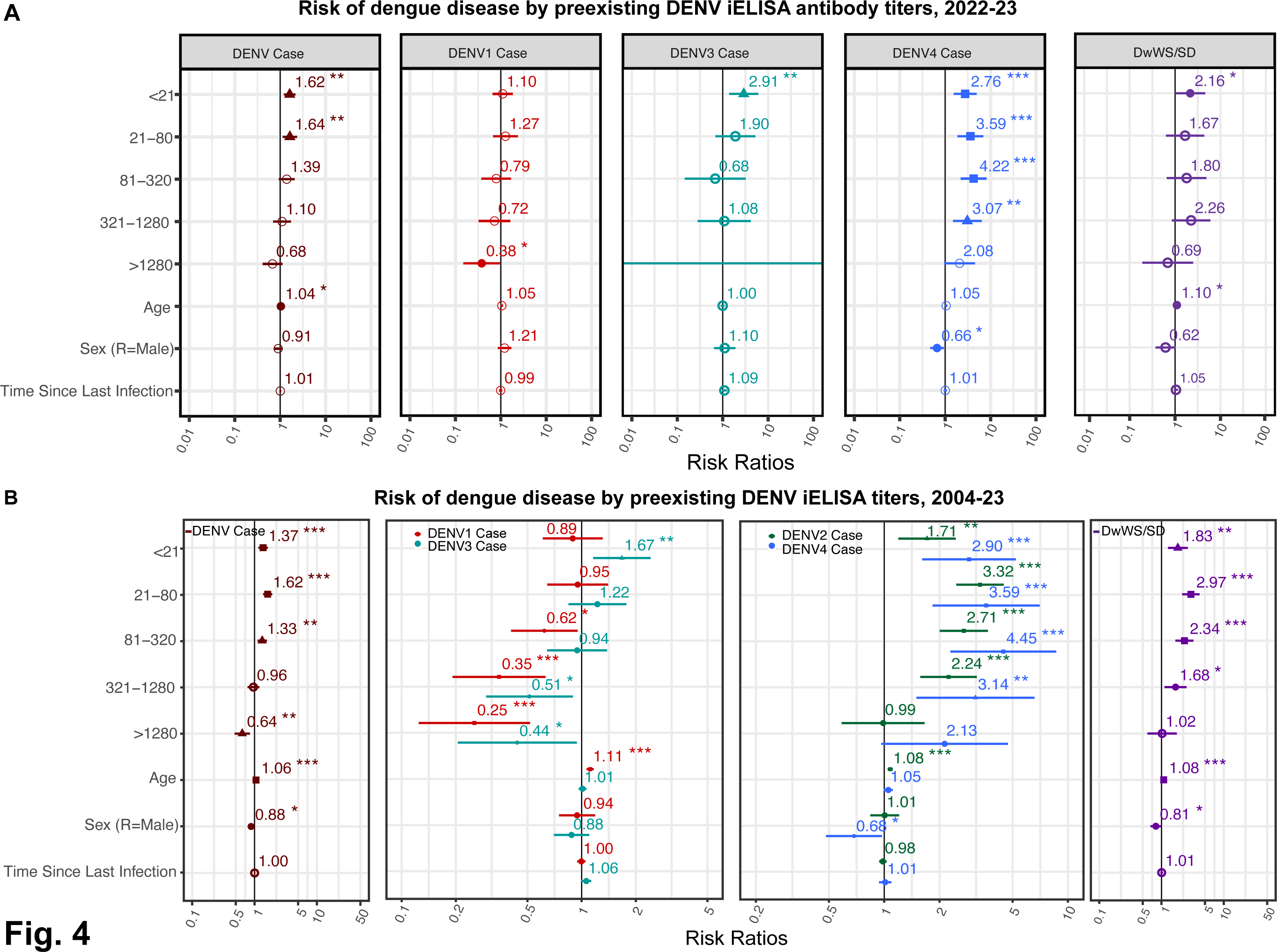
Risks of dengue differ by pre-existing anti-DENV antibody titers and infecting serotype. Linear mixed effects models were used to estimate the risk of any symptomatic DENV infection, as well as a symptomatic infection caused by DENV1-4 by pre-existing DENV iELISA titer during **(A)** the 2022-2023 epidemic or **(B)** all years of the cohort study (2004-2023).

### Antibody titers strongly explain the relationship between infection history and dengue risk

Based on our observations, we hypothesized that the effect of specific DENV and ZIKV infection histories on the risk of dengue is partially mediated (i.e., explained by) pre-existing DENV iELISA titers. Using a well-established approach, the mediation model, we tested the degree to which each infection history influences dengue disease outcome 1) directly, meaning the infection history itself affected outcome, versus 2) indirectly, meaning infection history modified antibody titers, which in turn modulated disease outcome. We analyzed the 2022-2023 epidemic season, and our models were adjusted for time since last infection, age, and sex. We separately tested the mediating effect of iELISA titers within a range previously identified as enhancing (10-320 vs. lower or higher titers) as opposed to protective (>320 vs. ≤320).

Consistently, every infection history induced antibodies in the range of 10-320 that were associated with increased risk of dengue (Fig. 5A). Titers between 10-320 explained a smaller fraction (<25%) of the total effect of DENV-DENV, ZIKV-DENV and 2+DENV infection histories on dengue outcome, while antibodies in this range explained a larger fraction of the effect of DENV, DENV-immune, ZIKV, DENV-ZIKV, and 2+DENV-ZIKV infection histories (Fig. 5A). Additionally, ZIKV infection had a direct effect (independent of titer) on increasing dengue risk, while the 2+DENV history had a direct protective effect. Conversely, when we tested whether titers greater than 320 (as compared to lower titers) mediated the relationship between infection history and dengue outcome, we found that high antibody titers showed a trend toward protection against symptomatic DENV infection, although this effect was not significant. High antibody titers explained almost all of the effect on dengue outcome for ZIKV-DENV history and almost none of the effect for the ZIKV group, while for other histories, titer explained about a quarter of the effect (Fig. 5B). ZIKV, DENV-DENV, and DENV-ZIKV still retained direct effects on increasing dengue risk. Notably, for infection histories that predisposed individuals to a greater risk of symptomatic DENV infection, such as DENV-DENV, high antibodies decreased their risk probabilities, although not significantly (Fig. 5B). On the contrary, for those categories with less risk, including 2+DENV or ZIKV-DENV, antibodies increased protection against symptomatic DENV infection (Fig. 5B).

**Fig. 5.**
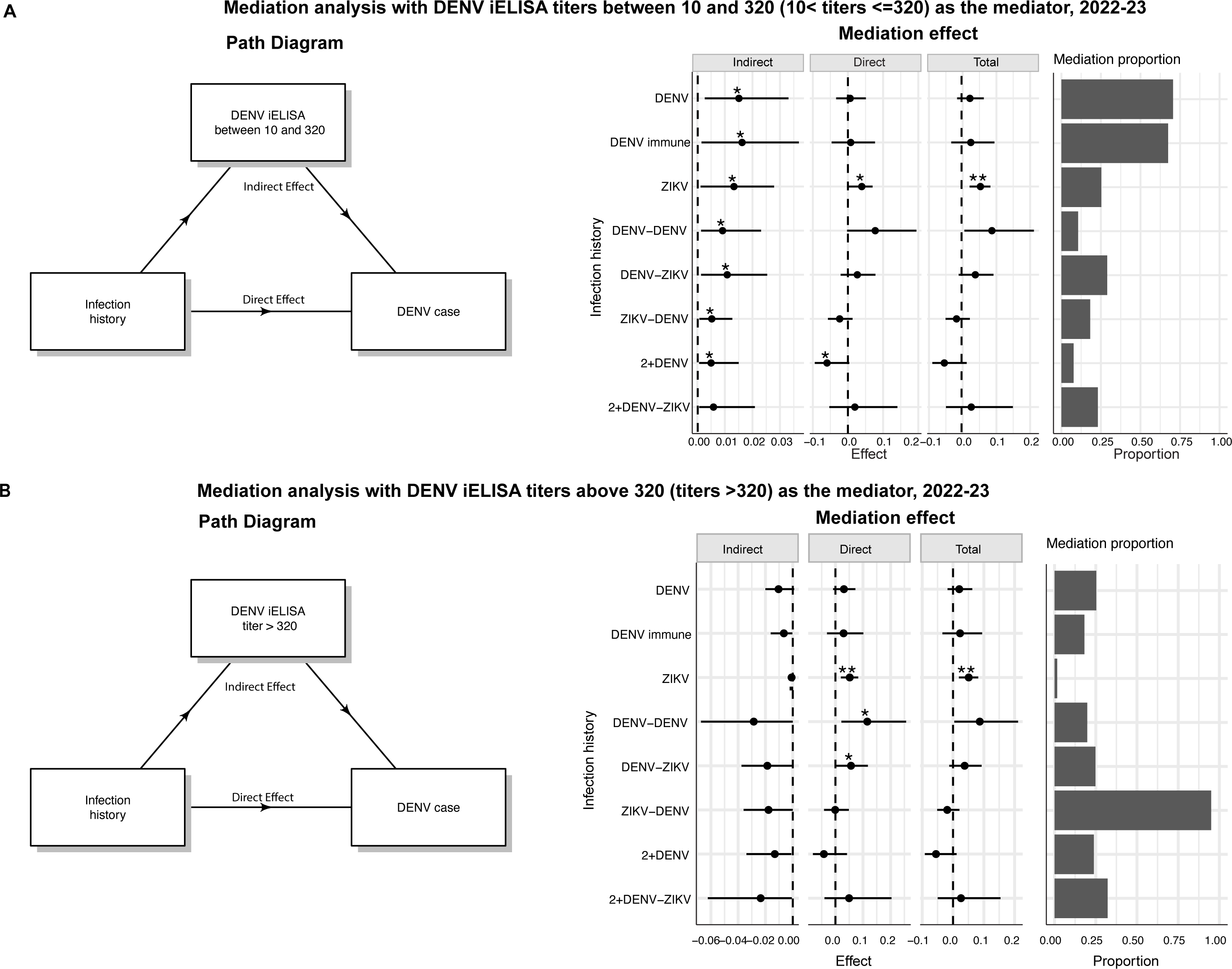
DENV iELISA titer is a mediator that helps explain the relationship between infection history and disease outcome. A) Mediation path diagram and effects and contribution of DENV iELISA titers (between 10 and 320) on disease risk attributed to prior flavivirus infection. **B)** Mediation path diagram and effects and contribution of DENV iELISA titers (>320) on disease risk attributed to prior flavivirus infection.

### Order of infecting DENV and ZIKV infections modifies antibody magnitude and kinetics

Cross-reactive anti-DENV antibody titers are modulated both by time since last infection and infection history. We have previously shown that anti-DENV antibodies measured by the DENV iELISA are stable or increase over time after a primary DENV and ZIKV infection and wane after secondary DENV or sequential DENV and ZIKV infections (*28*). Here, we characterized the magnitude and kinetics of anti-DENV antibodies in previously reported infection histories (*28*), and for the first time, in individuals with ZIKV-DENV infection histories. We noted differences in the magnitude of the iELISA antibody response measured in the annual sample after infection. Primary ZIKV infection induced the lowest antibody titers, followed by a primary DENV infection (Fig. 6 and table S6). Interestingly, the ZIKV-DENV group on average had 1.1- fold higher titers (p=3.2e-10) than the DENV-ZIKV group at the same timepoint (Fig. 6 and table S7), suggesting order of infection has a modest effect on the magnitude of cross-reactive anti-DENV antibodies. After a primary ZIKV infection, antibodies increased (half-life, t_1/2_: 2.31 years, 95% CI: 2.10-2.48) for three years, and then gradually declined (t_1/2_: 3.65 years, 3.01- 4.33), whereas after a primary DENV infections antibodies were stable (Fig. 6 and table S7). We also found that after sequential DENV or DENV and ZIKV infections, on average, antibodies initially wane rapidly in the first ∼2 years, with a subsequent gradual waning or stabilization. In particular, the ZIKV-DENV group was still experiencing rapid waning at the last timepoint measured, suggesting this infection history may be less protective long-term if their antibodies continue to wane.

**Fig. 6.**
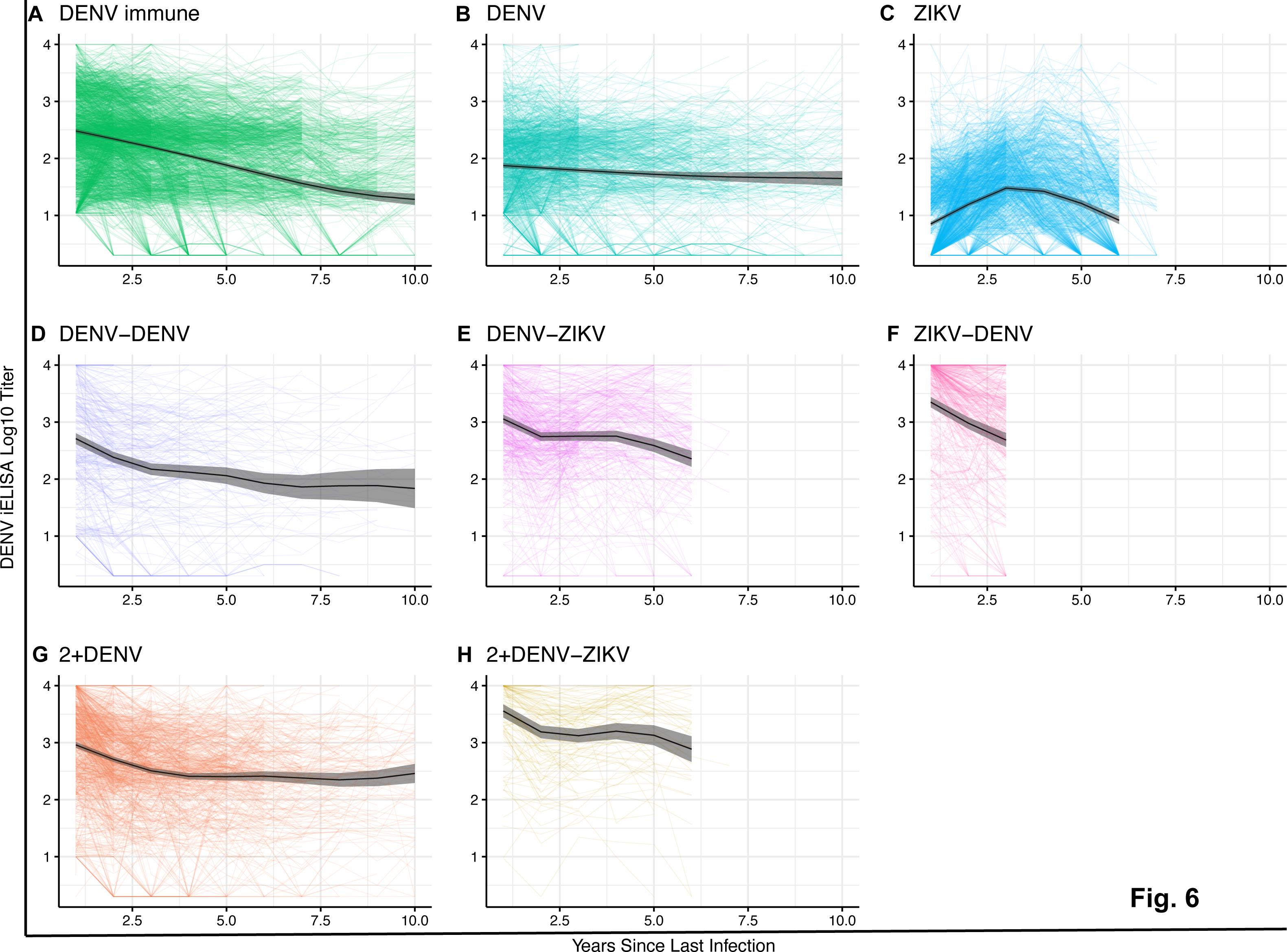
Primary DENV and ZIKV infections have distinct antibody dynamics as well as the order of DENV or ZIKV infections. Antibody kinetics in the Pediatric Dengue Cohort Study, as measured using the DENV iELISA, are shown. Data are shown for 1 to up to 10 years for the following infection histories: DENV-immune **(A),** primary DENV **(B)**, primary ZIKV **(C)**, DENV-DENV **(D**), DENV-ZIKV **(E),** ZIKV-DENV **(F)**, 2+DENV **(G),** and 2+DENV-ZIKV **(H)** infections. Individual trajectories and group Generalized Additive Model fits (black line; 95% CIs, gray shading) are shown.

## DISCUSSION

Our study presents novel insights into the complex interplay between the immune responses to DENV and ZIKV infections and their clinical and epidemiological outcomes. To our knowledge, this is the first study in humans to demonstrate that primary ZIKV infection increases risk of subsequent dengue disease in a serotype-dependent manner. We also show that DENV-ZIKV infection histories are associated with increased risk of symptomatic DENV2 and DENV4 infection, while ZIKV-DENV infection history is not. Pre-existing antibody titers were found to modulate disease risk depending on the incoming serotype. Finally, we show that most infection histories can elicit both enhancing and protective antibodies, and their relationship to DENV infection outcome is strongly dependent on antibody titer.

We have previously shown that a prior ZIKV infection increased risk of symptomatic and severe DENV2 (*26*). Here, using data from 374 cases in the 2022-2023 epidemic, we show that primary ZIKV infection also increased the risk of symptomatic DENV3 and DENV4 infections and overall disease severity. Intriguingly, a prior ZIKV infection was not a risk factor for symptomatic DENV1 infection. This matched our analysis showing that primary DENV infection is a risk factor for severe disease and symptomatic DENV2 and DENV4 infections across all cohort years. The enhancing effect of ZIKV has now been observed in two separate epidemics and against multiple DENV serotypes in Nicaragua. A separate study in Brazil of 879 dengue cases, >99% of which were caused by DENV2, similarly found that individuals with only prior ZIKV infection were more likely to experience severe disease and hospitalization than flavivirus-naïve participants (*17*). In this study, the authors found that the primary ZIKV group did not have higher viremia or pro-inflammatory cytokines than the DENV group, leading them to propose that the mechanism of enhanced severity may differ from classical ADE. Collectively, these observations suggest that induction of primary ZIKV immunity may increase risk of future dengue disease, posing a potential challenge to development of Zika vaccines.

Our study also shows that the order of sequential DENV and ZIKV infections modulates future disease outcome. DENV followed by ZIKV infection increased the risk of a symptomatic and severe DENV infection, whereas ZIKV followed by DENV infection did not. When stratified by incoming serotype, we found that DENV-ZIKV infection histories increased the risk only for DENV2 and DENV4, again showing immunological similarities in secondary DENV infections by these two viruses. A study in macaques has similarly found that DENV-ZIKV sequence produced an inferior immune response compared to ZIKV-DENV exposed monkeys, impacting the quality of antibodies produced in response to tertiary DENV infection and the magnitude of recall memory immune response, suggesting important differences by sequential order of infection (*19, 29, 30*). Notably, a study in Sri Lanka showed that individuals with prior DENV and ZIKV infection histories or only prior DENV infection experience increased risk of plasma leakage, shock, and severe dengue compared to flavivirus-naïve individuals, although order of infections could not be determined (*31*).

Interestingly, we found that having had two prior DENV infections was a significant risk factor for subsequent symptomatic and severe DENV infections as well as DENV3 symptomatic infection. However, we did not observe increased disease risk in groups with at least two prior DENV infections, with or without subsequent ZIKV infection (2+DENV and 2+DENV-ZIKV), and protective effects were observed for the 2+DENV group against any dengue case and symptomatic DENV1 infection. These groups include some individuals who entered the study with detectable anti-DENV antibodies and then experienced at least one DENV infection, meaning that some may have had only two prior infections. One interpretation of these findings is that there is something distinct about the DENV-DENV group, which constitutes a small fraction of the overall cohort. They may be at higher risk for exposure, although our neighborhood level analyses did not support this hypothesis. Alternatively, these findings may suggest that two DENV infections are not always be enough to provide broad protection against symptomatic DENV infection. Prior work has suggested that tertiary infections are rare in hospital settings (*32, 33*). Here we report mild-to-severe tertiary infections, as our community-based cohort allowed us to capture a greater spectrum of the disease. Our results are consistent with prior reports from the Pediatric Dengue Cohort Study that tertiary infections for some epidemic years were more likely to be symptomatic than inapparent (*34*). Although we previously found that having two or more DENV infections protected against DENV2 cases during the 2019-2020 epidemic, DENV2 had circulated in the years before that epidemic, implying that one of the prior infections may have been with DENV2 and thus that homotypic immunity contributed to reduced risk (*35, 36*). In contrast, DENV1 and DENV3 have not circulated appreciably in Nicaragua for more than 10 years, and our pediatric study population had never experienced a large DENV4 epidemic. Lack of homotypic immunity against these serotypes and insufficient heterotypic immunity may have driven the explosive dengue epidemic in 2022-2023 and explained the lack of protection even in those with two prior DENV infections.

Our findings also support the previous observation that pre-existing DENV immunity differentially modulates subsequent symptomatic and severe disease depending on infecting serotype. We previously showed that low-to-intermediate pre-existing DENV iELISA antibody titers were associated with increased symptomatic and severe DENV2 and DENV3 disease, whereas high titers were protective against DENV1 and DENV3 (*26*). Here, we found that a history of any flavivirus infection increased the risk of future DwWS/SD and DHF/DSS as well as dengue cases caused by DENV2 and DENV4, but was protective against DENV1 and had neutral risk against DENV3. Low-to-intermediate pre-existing antibody titers increased risk of DENV2 and DENV4 infections compared to flavivirus-naïve individuals, and high titers were not associated with increased risk nor protection. In contrast, high pre-existing antibodies reduced risk of symptomatic DENV1 and DENV3 infections, while low titers were associated with increased risk of symptomatic DENV3 infection. Structural differences between the viruses may explain a lower probability of ADE with DENV1, which is associated with lower frequency of symptomatic and severe secondary infections (*37*). DENV1 has been shown to produce more mature viral particles *in vivo*, which are poorly neutralized by fusion loop antibodies, and it has been shown *in vitro* that some DENV1 particles breathe less compared to other serotypes (*38, 39*).

We also showed that anti-DENV antibodies help mediate the relationship between DENV or ZIKV infection and risk of subsequent symptomatic DENV infection via antibodies. Across infection histories, we found that DENV iELISA antibody titers are good predictors of disease outcome. This observation is consistent with previous studies showing that high anti-DENV antibody titers are good correlates of protection following vaccination with dengue candidate CYD-TDV, while a lower titer was a correlate of risk (*40*). Notably, some infection histories still retain direct effects, suggesting that there are differences in the quality of immunity that are not fully captured by our measure of anti-DENV antibodies.

Prior work has suggested that cross-reactive protection against DENV wanes within one to three years post-infection, and a longer time between sequential DENV infections increases the probability of symptomatic DENV infections (*32, 34, 41, 42*). Our previous work showed that cross-reactive antibodies after a primary DENV infection are stable by about 6 months, while primary ZIKV infection elicits cross-reactive DENV antibodies that increased over time up to three years (*28*). In contrast, after secondary DENV or DENV-ZIKV infection, we observed high antibodies followed by long-term decay. Here, we show that after 3 years, DENV cross-reactive antibodies in primary ZIKV participants begin to decay. We also showed that the ZIKV-DENV group elicited slightly higher DENV iELISA titer than the DENV-ZIKV group did, suggesting broader immunity, which could have resulted in a reduction of risk for a subsequent symptomatic DENV infection. However, the ZIKV-DENV group experienced rapid antibody decay, raising the possibility that the protective effect may not be long-lived.

Our study has several strengths, including a robust study design that has been sustained over 19 years. This extensive period of observation has enabled us to detect epidemics of all DENV serotypes and ZIKV, encompassing the entire spectrum of disease severity, from inapparent to symptomatic and severe infections. This comprehensive dataset allows the capture of detailed arboviral infection histories, including single and sequential DENV and ZIKV infections, with a large sample size. Nonetheless, our study has several limitations. The data is derived from a Nicaraguan cohort, and the results may not generalize to other settings with different DENV and ZIKV strains as well as other circulating flaviviruses. However, given the similarity in dengue epidemiology across the Americas, our findings will likely have broader applicability in the region (*43–45*). Additionally, the exact sequence of infecting DENV serotypes is not recorded due to a lack of serotype information of inapparent infections. The sequence of infecting DENV serotypes may modulate disease outcome in ways we could not account for here. Further, risk calculations using all Pediatric Dengue Cohort Study participants in the denominator assumes risk is homogenous, which is implausible. To account for this, we adjusted results by prior year DENV seroprevalence at the neighborhood levels as a proxy for spatial risk and showed that our results did not qualitatively change. Finally, our mediation analysis should not be interpreted causally as it relies on diverse assumptions that cannot be fully satisfied using our study design.

In summary, this study highlights the serotype-specific modulation of infection risk based on prior flavivirus exposure. The observation that individuals with prior ZIKV, DENV, or DENV-ZIKV infections were at increased risk of symptomatic dengue caused by multiple DENV serotypes suggests that ADE potential is not limited to DENV2 but also extends DENV3 and DENV4. The findings presented here, in conjunction with other studies, suggest that the introduction of a monovalent ZIKV vaccine that induces immunity similar to natural ZIKV infection could have a negative effect on dengue outcome if introduced to DENV-naïve populations. These findings advocate a more nuanced and integrative approach to vaccine deployment strategies, considering the history of flavivirus infections and the level of flavivirus immunity in the target population. Further, our results suggest that introducing ZIKV vaccines is unlikely to broaden protective immunity in individuals with one prior DENV infection, and if anything, may increase future dengue risk. Our findings also suggest that some individuals with two natural DENV infections remain at risk of DwWS/SD and may suggest that even some individuals who are DENV immune before receiving dengue vaccines may remain at heightened risk for disease. The possibility that vaccine-induced immunity could inadvertently elevate the risk of severe disease warrants an evidence-based approach to vaccination programs and longer-term follow-up times to evaluate protection against all DENV serotypes.

## MATERIALS AND METHODS

### Study design

This study aimed to assess how pre-existing ZIKV and DENV immunity impacts the risk of symptomatic DENV1-4 and severe dengue disease, at a population level. To do this, we analyzed serological data collected since 2004 from a continuous observational cohort study in Nicaragua, adhering to the guidelines in the Strengthening the Reporting of Observational Studies in Epidemiology checklist. This study did not involve any form of randomization or blinding measures. Our analysis included all individuals, with results reported only for those with DENV and ZIKV infection histories that could be clearly determined. All relevant data available during the manuscript preparation was included. The primary endpoint was the risk of symptomatic DENV infection influenced by previous ZIKV or DENV immunity and the infecting DENV serotype. Secondary endpoints were the impact of pre-existing antibody titers on dengue and severe dengue risk, as well as antibody magnitude and long-term kinetics following exposure to one or multiple flaviviruses.

### Ethics statement

The human subjects protocol of the Pediatric Dengue Cohort Study was reviewed and approved by the Institutional Review Boards of the University of California, Berkeley (2010-09-2245), the University of Michigan (HUM00091606), and the Nicaraguan Ministry of Health (CIRE- 09/03/07-008). Parents or legal guardians of all subjects provided written informed consent, and participants aged 6 years of age and older provided assent.

### The Pediatric Dengue Cohort Study

The Pediatric Dengue Cohort Study is an ongoing prospective study that began in 2004 and is conducted at the Sócrates Flores Vivas Health Center (HCSFV), based in District 2 of Managua, Nicaragua (*46*). This district is adjacent to Lake Managua and consists of neighborhoods ranging from low to middle socioeconomic status (SES). The HCSFV serves approximately 62,000 residents across 18 neighborhoods within District 2 (*47*). The illiteracy rate in this area is approximately 7%, and over 90% of the households have access to running water and a sewage system. The streets are largely paved, and all residents have access to electricity (*48*).

The study tracks approximately 4,000 active participants yearly and has followed 9,800 children since its inception. To maintain the original age distribution of the cohort, ∼300 two-year-olds, along with some children aged 3-11 years, are enrolled in the study each year. The age limit of the study was increased to 15, 16, and 17 years in 2018, 2019, and 2022, respectively. Participants are provided free medical care 24 hours per day, 365 days per year, through study physicians at the HCSFV. If a child requires hospitalization, they are transferred by study staff to the study hospital, the National Pediatric Reference Hospital (Hospital Infantil Manuel de Jesús Rivera). Children presenting with fever and since July 2016, rash, are screened for signs and symptoms of dengue, Zika, and chikungunya. Symptomatic infections are captured through passive surveillance, enhanced with periodic home visits. Acute (0-5 days post symptoms onset) and convalescent samples (14-21 days) are collected for each suspected case to confirm infection (*35*). Every year in March-April (previously in July before 2011), healthy blood samples are provided by all participants to evaluate inapparent arboviral infections (*35*).

### Case definitions

Upon medical examination, children are categorized into A, B, C, or D cases, as follows: **A**: those meeting the 1997 or 2009 World Health Organization (WHO) criteria for dengue; **B**: those with undifferentiated fever of undefined origin; **C**: those with an acute febrile illness identified as a disease other than dengue, chikungunya, or Zika; **D**: those with other conditions (including rash) that do not include a fever. A range of clinical and laboratory information is collected for each participant, covering 189 different variables (*35*), including vital signs, temperature, musculoskeletal pain, respiratory symptoms, gastrointestinal symptoms, indicators of dehydration, rashes, and other skin anomalies. Since 2004, all cases falling into categories A and B are tested for acute DENV infection and, starting in 2014, also for chikungunya virus (CHIKV). As of 2016, all cases meeting the definitions of A, B, or D (rash without fever) are tested for acute DENV, CHIKV, and ZIKV infection.

### Detection of cases

Acute samples are examined using the ZCD multiplex real-time reverse transcriptase PCR (rRT-PCR) assay for DENV, ZIKV, and CHIKV (*49*); prior to the introduction of ZIKV into the cohort, other RT-PCR methods were used (*50–52*). DENV-positive samples are further analyzed by rRT-PCR to determine the infecting DENV serotype, and a subset undergo virus isolation in C6/36 cells (*50, 53*). Paired acute-phase (days 1-5) and convalescent-phase (days 14-21) samples are evaluated for seroconversion of IgM antibodies, an indicator of a recent infection, using separate in-house DENV and ZIKV IgM capture (MAC)- ELISAs (*54, 55*). DENV inhibition ELISA (iELISA) and ZIKV iELISA assays are used to measure total anti-DENV and -ZIKV antibodies, respectively, and to detect seroconversion or a 4-fold or greater increase in antibodies (*12, 54*). For arbovirus cases from 2016-2017, we also used a computer algorithm to help identify Zika cases from dengue cases based on the results of the aforementioned antibody tests (*56*).

### Detection of inapparent infections

To detect inapparent DENV infections, before 2016, we identified an inapparent DENV infection if a seroconversion or a ≥4-fold increase in antibody titer was recorded between annual samples as measured by DENV iELISA, provided no observed febrile episode was identified as a dengue case (*12*). After the 2016 Zika epidemic, we defined inapparent DENV infections as seroconversion or a ≥4-fold rise in DENV iELISA titer without seroconversion by ZIKV NS1 BOB ELISA, seroconversion or a ≥4-fold rise by ZIKV iELISA, or an observed dengue, Zika, or flavivirus case in that year (*54*). Inapparent flavivirus (DENV or ZIKV without ascertainment) infections were defined by seroconversion or a ≥4-fold rise by both DENV iELISA and ZIKV iELISA, no seroconversion by ZIKV NS1 BOB ELISA, and no documented dengue, Zika, or flavivirus case in that year. Seroconversion by ZIKV NS1 BOB ELISA and/or seroconversion or ≥4-fold rise by ZIKV iELISA was used to detect inapparent ZIKV infections.

### Infection histories

The study participants were grouped into categories based on their DENV and/or ZIKV infection history. When a child experienced another infection, they were moved to the group that reflected their new infection status. We note that none of the groups with ZIKV existed before the major Zika epidemic in 2016. The different groups included children who: were free from DENV (from 2004-2015) or ZIKV (from 2016-2023) immunity at the start of the study and did not experience a DENV, ZIKV, or flavivirus infection during the study period **(flavivirus-naïve)**; entered the study with DENV immunity, determined using the DENV iELISA (titer ≥1:10) and with no antibody titers as measured in other assays or with low ZIKV titers and no subsequent detected infection **(DENV-immune)**; were initially free from flaviviruses but were observed to have experienced precisely one DENV infection **(DENV)**; were initially free from flaviviruses but who experienced exactly two documented DENV infections **(DENV-DENV)**. Since 2020, ZIKV titers were disregarded for new enrollees, given that most new enrollees did not experience the ZIKV epidemic, given their age, and there is no evidence that ZIKV has circulated after 2017. Other groups included children who: either entered the study without flavivirus immunity and experienced more than two documented DENV infections or who entered the study with flavivirus immunity and had one or more observed DENV infection **(2+DENV)**; experienced only one prior ZIKV infection **(ZIKV)** – these children either entered the study without flavivirus immunity and had one observed ZIKV infection or a flavivirus infection (see definition above; based on the epidemiology at the time, these are most likely ZIKV infections), or positive for ZIKV but with low DENV titers. The **DENV-ZIKV** group included children who experienced one DENV infection followed by one ZIKV infection; the **2+DENV-ZIKV** group included children with two or more prior DENV infections followed by one ZIKV infection; and the **ZIKV-DENV** group comprised children with a primary ZIKV infection followed by one DENV infection. The ZIKV-DENV group did not exist prior to the DENV2 epidemic in 2019. Other infection histories included children who had a ZIKV-DENV infection history who subsequently experienced another DENV infection (**ZIKV-DENV-DENV**); this group (N = 15) experienced no subsequent DENV infections and was not included in our analyses of infection histories, but they were included in the titer analyses. The **DENV-ZIKV-2+DENV** group consisted of participants who initially had a DENV-ZIKV-DENV infection history and later experienced another DENV infection. This group (N = 3) experienced no subsequent DENV infections and was not included in our analyses of infection histories, but they were included in the titer analyses. Finally, the **2+DENV-ZIKV-DENV** group consisted of participants who initially were in the 2+DENV group and later had one ZIKV and other DENV infection. Again, this group (N = 11) had no further infections and was not included in our analyses of infection histories but was included in the titer analyses. Some children with ZIKV infections in 2016- 2017 were observed to have increasing titers in DENV and ZIKV serological assays for multiple years. These infections were not considered actual infections and were therefore excluded when defining infection histories.

### Severity classification

Dengue cases in the study were classified according to the World Health Organization’s (WHO) guidelines from 1997 and 2009. According to the 1997 WHO case classification, Dengue Fever (DF) was defined as a febrile case with at least two of the following symptoms: headache, arthralgia, retro-orbital pain, rash, myalgia, hemorrhagic manifestations, and leukopenia. Dengue Hemorrhagic Fever (**DHF)** was defined as a laboratory-confirmed dengue case presenting with: 1) fever or a history of fever; 2) hemorrhagic manifestations; 3) thrombocytopenia (≤100,000 platelets/ml); and 4) signs of plasma leakage. The criteria for plasma leakage included age and sex-specific elevated hematocrit levels, ≥20% hemoconcentration, pleural effusion, ascites, edema, hypoproteinemia, hypoalbuminemia, or other evidence of plasma leak. Dengue Shock Syndrome (**DSS)** is a DHF case with signs of circulatory failure, including 1) hypotension for the patient’s age or a narrow pulse pressure (<20 mm Hg), and 2) clinical signs of shock such as a rapid, weak pulse, cold and clammy skin, or poor capillary refill.

According to the 2009 WHO case classification, Dengue with Warning Signs (DwWS) was used when a laboratory-confirmed case presented with symptoms such as abdominal pain, persistent vomiting, fluid accumulation, lethargy, mucosal bleeding, liver enlargement, or a rise in hematocrit concurrent with a decrease in platelet count. Severe Dengue (SD) indicated laboratory-confirmed cases with severe bleeding, significant plasma leakage leading to shock, fluid accumulation resulting in respiratory distress, or organ failure. Organ involvement was signified by liver ALT or AST levels of ≥1,000, impaired consciousness, or heart or other organ failures.

### Inclusion criteria for cases in the 2022 epidemic cases

Between March 19, 2022, and July 2, 2023, we documented 384 dengue cases. Notably, 15 of these cases were identified based on DENV serology, but they also exhibited cross-reactivity with ZIKV serology. Because ZIKV has not been detected since its initial introduction in the region in 2016, these cases were classified as DENV cases in the present study. We excluded 10 cases that had a second-laboratory-confirmed DENV infection in the same epidemic, including 8 where both infections were confirmed by RT-PCR, and 2 where one of the infections was confirmed by DENV serology. Our analysis ultimately utilized 374 total cases, including 39 identified through DENV serology.

### Statistical analyses

All statistical and epidemiological analyses were conducted using R (R Foundation for Statistical Computing, version 4.5.0) and used lme4, segmented, and mediation packages (*57–60*). Log-binomial and log-poisson generalized linear regression models (GLMs) were used to estimate the risk of any dengue case given infection history. Log-poisson GLMs were used when log-binomial models would not converge due to rare disease outcomes; that is, in instances where we had low numbers of dengue cases. Models were adjusted for age, sex, and time since last infection. All analyses were also performed separately for each infecting DENV serotype. We were unable to analyze DENV serotype in analyses of DENV severity due to sample size limitations. We calculated 95% confidence intervals (CIs) of the predicted values based on the standard errors (SE), assuming a normal distribution (predicted mean plus or minus ±1.96 multiplied by the SE). GLMs were used for all 2022-2023 epidemic analyses since each participant contributed only one year to the data.

For analyses using all 19 years of cohort data, generalized linear mixed models (GLMMs) were used to incorporate random effects, allowing us to account for repeated measurements by clustering at the individual level (longitudinal data analysis). We performed two main analyses: one that studied the effect of infection histories on DENV outcomes, and one that categorized participants as flavivirus-naïve versus those exposed to any number of DENV or ZIKV infections (flavi-immune). Additionally, we adjusted for age, sex, time since last infection, and cohort epidemic year. Neighborhood seroprevalence was also included as a proxy for exposure in sensitivity analyses. Log-binomial and log-poisson GLMMs were used to model the relationship between pre-existing DENV iELISA titers and disease outcome. These models were also adjusted for age, sex, time since the last infection and cohort epidemic year.

Mediation analysis was used to differentiate the direct versus indirect effect of antibody titers in explaining the relationship between infection history and disease presentation for the 2022-2023 epidemic. Using logistic regression models, adjusting for age, sex, and year, we first regressed the relationship between infection history and DENV iELISA titers and then we modeled the likelihood of being a DENV case as a function of infection history and DENV iELISA titers. We used non-parametric bootstrapping and bias-corrected, accelerated confidence intervals employing 1,000 simulations. The indirect effects were calculated as the expected difference in the potential outcomes if DENV iELISA titers were within a specific range versus outside of that range. We performed separate analyses comparing titers of 10-320 vs. higher or lower and >320 vs. ≤320. We performed pairwise comparisons of flavivirus-naïve vs non-flavivirus-naïve categories (DENV immune, DENV, ZIKV, DENV-DENV, DENV-ZIKV, ZIKV-DENV, 2+DENV, 2+DENV-ZIKV) resulting in eight comparisons or contrasts. The proportion of mediation was determined by dividing the indirect effect by the total effect. When a mediation model was inconsistent, meaning that the direct and indirect effects were in opposite signs, the absolute value of them were used to calculate the mediation contribution proportion (*61*).

We measured antibody dynamics by infection history group using Gaussian generalized additive mixed models (GAMMs) that included random effects for both the intercept and slope, as each individual had a different starting titer magnitude and titer kinetics. Antibody dynamics analyses were adjusted for age and sex. For computational efficiency purposes, in order to account for the individual variability, all continuous covariates were discretized, including time since last infection (but allowing time since last infection to be smooth) and variance parameters were estimated using a fast restricted maximun likelihood (REML) function (*62*). We used these models to identify relevant inflection points in the antibody kinetics for further analyses related to intercepts and slopes within each infection history group. Multiphasic linear models were then used to model the various phases of antibody decay or rise while estimating antibody half-lives. Half-lives were calculated by dividing log(2) titers by the slope. A positive half-life indicated that antibody titers waned, whereas a negative-half-life represented increasing antibody titers.

## Data Availability

All data pertinent to this study can be found within the paper or the Supplementary Materials. and the associated code is available for reference on Zenodo. After securing approval from the UC Berkeley Committee for the Protection of Human Subjects, individual data for figure reproduction could be shared with external researchers. For data access arrangements, please contact EH at. Standard data and material transfer agreements govern all materials and data used in this study.

## Acknowledgments

We thank the participants of the Pediatric Dengue Cohort Study and their families. We are grateful to both past and current team members of the study, based at the Sócrates Flores Vivas Health Center, the Laboratorio Nacional de Virología in the Centro Nacional de Diagnóstico y Referencia, and the Sustainable Sciences Institute in Nicaragua for their commitment and exceptional work.

## Author contributions

Conceptualization: JVZ, CMH, RAA, SB, LCK, EH

Methodology: JVZ, CMH, RAA, LCK, EH

Investigation: SA, CN, KG, DC, TM, GK, AB

Visualization: JVZ, CMH, RAA

Funding acquisition: AB, LCK, EH

Project administration: GK, AG, AB, EH

Supervision: LCK, EH

Writing – original draft: JVZ, CMH, RAA, LCK, EH

Writing – review & editing: All authors.

## Supplementary materials

Fig. S1 to S3 Table S1 to S7

## References

1. S. Leta, T. J. Beyene, E. M. De Clercq, K. Amenu, M. U. G. Kraemer, C. W. Revie, Global risk mapping for major diseases transmitted by Aedes aegypti and Aedes albopictus. Int. J. Infect. Dis. 67, 25–35 (2018).

2. L. Cattarino, I. Rodriguez-Barraquer, N. Imai, D. A. T. Cummings, N. M. Ferguson, Mapping global variation in dengue transmission intensity. Sci. Transl. Med. 12, eaax4144 (2020).

3. N. R. Faria, J. Quick, I. M. Claro, J. Theze, J. G. de Jesus, M. Giovanetti, M. U. G. Kraemer, S. C. Hill, A. Black, A. C. da Costa, L. C. Franco, S. P. Silva, C. H. Wu, J. Raghwani, S. Cauchemez, L. du Plessis, M. P. Verotti, W. K. de Oliveira, E. H. Carmo, G. E. Coelho, Acfs Santelli, L. C. Vinhal, C. M. Henriques, J. T. Simpson, M. Loose, K. G. Andersen, N. D. Grubaugh, S. Somasekar, C. Y. Chiu, J. E. Munoz-Medina, C. R. Gonzalez-Bonilla, C. F. Arias, L. L. Lewis-Ximenez, S. A. Baylis, A. O. Chieppe, S. F. Aguiar, C. A. Fernandes, P. S. Lemos, B. L. S. Nascimento, H. A. O. Monteiro, I. C. Siqueira, M. G. de Queiroz, T. R. de Souza, J. F. Bezerra, M. R. Lemos, G. F. Pereira, D. Loudal, L. C. Moura, R. Dhalia, R. F. Franca, T. Magalhaes, E. T. Marques, Jr., T. Jaenisch, G. L. Wallau, M. C. de Lima, V. Nascimento, E. M. de Cerqueira, M. M. de Lima, D. L. Mascarenhas, J. P. M. Neto, A. S. Levin, T. R. Tozetto-Mendoza, S. N. Fonseca, M. C. Mendes-Correa, F. P. Milagres, A. Segurado, E. C. Holmes, A. Rambaut, T. Bedford, M. R. T. Nunes, E. C. Sabino, L. C. J. Alcantara, N. J. Loman, O. G. Pybus, Establishment and cryptic transmission of Zika virus in Brazil and the Americas. Nature 546, 406–410 (2017).

4. A. E. Ngono, S. Shresta, Immune Response to Dengue and Zika. Annu. Rev. Immunol. 36, 279–308 (2018).

5. L. C. Katzelnick, S. Bos, E. Harris, Protective and enhancing interactions among dengue viruses 1-4 and Zika virus. Curr. Opin. Virol. 43, 59–70 (2020).

6. E. N. Gallichotte, R. S. Baric, A. M. de Silva, The Molecular Specificity of the Human Antibody Response to Dengue Virus Infections. Adv. Exp. Med. Biol. 1062, 63–76 (2018).

7. L. C. Katzelnick, M. Montoya, L. Gresh, A. Balmaseda, E. Harris, Neutralizing antibody titers against dengue virus correlate with protection from symptomatic infection in a longitudinal cohort. Proc. Natl. Acad. Sci. U S A 113, 728–733 (2016).

8. S. Bos, A. L. Graber, J. A. Cardona-Ospina, E. M. Duarte, J. V. Zambrana, J. A. Ruiz Salinas, R. Mercado-Hernandez, T. Singh, L. C. Katzelnick, A. de Silva, G. Kuan, A. Balmaseda, E. Harris, The association of neutralizing antibodies with protection against symptomatic dengue virus infection varies by serotype, prior infection history, and assay condition. *medRxiv*, Preprint at: 10.1101/2023.06.20.23291522 (2023).

9. D. Buddhari, J. Aldstadt, T. P. Endy, A. Srikiatkhachorn, B. Thaisomboonsuk, C. Klungthong, A. Nisalak, B. Khuntirat, R. G. Jarman, S. Fernandez, S. J. Thomas, T. W. Scott, A. L. Rothman, I. K. Yoon, Dengue virus neutralizing antibody levels associated with protection from infection in thai cluster studies. PLoS Negl. Trop. Dis. 8, e3230 (2014).

10. A. Nisalak, H. E. Clapham, S. Kalayanarooj, C. Klungthong, B. Thaisomboonsuk, S. Fernandez, J. Reiser, A. Srikiatkhachorn, L. R. Macareo, J. T. Lessler, D. A. Cummings, I. K. Yoon, Forty Years of Dengue Surveillance at a Tertiary Pediatric Hospital in Bangkok, Thailand, 1973-2012. Am. J. Trop. Med. Hyg. 94, 1342-1347 (2016).

11. N. Sangkawibha, S. Rojanasuphot, S. Ahandrik, S. Viriyapongse, S. Jatanasen, V. Salitul, B. Phanthumachinda, S. B. Halstead, Risk factors in dengue shock syndrome: a prospective epidemiologic study in Rayong, Thailand. I. The 1980 outbreak. Am. J. Epidemiol. 120, 653–669 (1984).

12. L. C. Katzelnick, L. Gresh, M. E. Halloran, J. C. Mercado, G. Kuan, A. Gordon, A. Balmaseda, E. Harris, Antibody-dependent enhancement of severe dengue disease in humans. Science 358, 929–932 (2017).

13. H. Salje, D. A. T. Cummings, I. Rodriguez-Barraquer, L. C. Katzelnick, J. Lessler, C. Klungthong, B. Thaisomboonsuk, A. Nisalak, A. Weg, D. Ellison, L. Macareo, I. K. Yoon, R. Jarman, S. Thomas, A. L. Rothman, T. Endy, S. Cauchemez, Reconstruction of antibody dynamics and infection histories to evaluate dengue risk. Nature 557, 719–723 (2018).

14. S. B. Halstead, S. Nimmannitya, S. N. Cohen, Observations related to pathogenesis of dengue hemorrhagic fever. IV. Relation of disease severity to antibody response and virus recovered. Yale. J. Biol. Med. 42, 311–328 (1970).

15. S. B. Halstead, E. J. O’Rourke, Dengue viruses and mononuclear phagocytes. I. Infection enhancement by non-neutralizing antibody. J. Exp. Med. 146, 201–217 (1977).

16. M. G. Guzman, M. Alvarez, S. B. Halstead, Secondary infection as a risk factor for dengue hemorrhagic fever/dengue shock syndrome: an historical perspective and role of antibody-dependent enhancement of infection. Arch. Virol. 158, 1445–1459 (2013).

17. C. F. Estofolete, A. F. Versiani, F. S. Dourado, Bhga Milhim, C. C. Pacca, G. C. D. Silva, N. Zini, B. F. D. Santos, F. A. Gandolfi, N. F. B. Mistrao, P. H. C. Garcia, R. S. Rocha, L. Gehrke, I. Bosch, R. E. Marques, M. M. Teixeira, F. G. da Fonseca, N. Vasilakis, M. L. Nogueira, Influence of previous Zika virus infection on acute dengue episode. PLoS Negl. Trop. Dis. 17, e0011710 (2023).

18. Katzelnick LC, Narvaez C, Arguello S, Lopez Mercado B, Collado D, Ampie O, Elizondo D, Miranda T, Bustos Carillo F, Mercado JC, Latta K, Schiller A, Segovia-Chumbez B, Ojeda S, Sanchez N, Plazaola M, Coloma J, Halloran ME, Premkumar L, Gordon A, Narvaez F, de Silva AM, Kuan G, Balmaseda A, Harris E, Zika virus infection enhances future risk of severe dengue disease. *Science (New York*, N.Y.) 369, 1123–1128 (2020).

19. M. E. Breitbach, C. M. Newman, D. M. Dudley, L. M. Stewart, M. T. Aliota, M. R. Koenig, P. M. Shepherd, K. Yamamoto, C. M. Crooks, G. Young, M. R. Semler, A. M. Weiler, G. L. Barry, H. Heimsath, E. L. Mohr, J. Eichkoff, W. Newton, E. Peterson, N. Schultz-Darken, S. R. Permar, H. Dean, S. Capuano, 3rd, J. E. Osorio, T. C. Friedrich, D. H. O’Connor, Primary infection with dengue or Zika virus does not affect the severity of heterologous secondary infection in macaques. PLoS Pathog. 15, e1007766 (2019).

20. J. George, W. G. Valiant, M. J. Mattapallil, M. Walker, Y. S. Huang, D. L. Vanlandingham, J. Misamore, J. Greenhouse, D. E. Weiss, D. Verthelyi, S. Higgs, H. Andersen, M. G. Lewis, J. J. Mattapallil, Prior Exposure to Zika Virus Significantly Enhances Peak Dengue-2 Viremia in Rhesus Macaques. Sci Rep 7, 10498 (2017).

21. M. G. Guzman, G. Kouri, L. Valdes, J. Bravo, M. Alvarez, S. Vazques, I. Delgado, S. B. Halstead, Epidemiologic studies on Dengue in Santiago de Cuba, 1997. Am. J. Epidemiol. 152, 793-799 (2000).

22. I. Y. Amaya-Larios, R. A. Martinez-Vega, F. A. Diaz-Quijano, E. Sarti, E. Puentes- Rosas, L. Chihu, J. Ramos-Castaneda, Risk of dengue virus infection according to serostatus in individuals from dengue endemic areas of Mexico. Sci Rep 10, 19017 (2020).

23. S. Henein, J. Swanstrom, A. M. Byers, J. M. Moser, S. F. Shaik, M. Bonaparte, N. Jackson, B. Guy, R. Baric, A. M. de Silva, Dissecting Antibodies Induced by a Chimeric Yellow Fever-Dengue, Live-Attenuated, Tetravalent Dengue Vaccine (CYD-TDV) in Naive and Dengue-Exposed Individuals. J. Infect. Dis. 215, 351–358 (2017).

24. E. Harris, E. Videa, L. Perez, E. Sandoval, Y. Tellez, M. L. Perez, R. Cuadra, J. Rocha, W. Idiaquez, R. E. Alonso, M. A. Delgado, L. A. Campo, F. Acevedo, A. Gonzalez, J. J. Amador, A. Balmaseda, Clinical, epidemiologic, and virologic features of dengue in the 1998 epidemic in Nicaragua. Am. J. Trop. Med. Hyg. 63, 5–11 (2000).

25. Z. Moodie, M. Juraska, Y. Huang, Y. Zhuang, Y. Fong, L. N. Carpp, S. G. Self, L. Chambonneau, R. Small, N. Jackson, F. Noriega, P. B. Gilbert, Neutralizing Antibody Correlates Analysis of Tetravalent Dengue Vaccine Efficacy Trials in Asia and Latin America. J. Infect. Dis. 217, 742–753 (2018).

26. L. C. Katzelnick, C. Narvaez, S. Arguello, B. Lopez Mercado, D. Collado, O. Ampie, D. Elizondo, T. Miranda, F. Bustos Carillo, J. C. Mercado, K. Latta, A. Schiller, B. Segovia-Chumbez, S. Ojeda, N. Sanchez, M. Plazaola, J. Coloma, M. E. Halloran, L. Premkumar, A. Gordon, F. Narvaez, A. M. de Silva, G. Kuan, A. Balmaseda, E. Harris, Zika virus infection enhances future risk of severe dengue disease. Science 369, 1123–1128 (2020).

27. S. Olkowski, B. M. Forshey, A. C. Morrison, C. Rocha, S. Vilcarromero, E. S. Halsey, T. J. Kochel, T. W. Scott, S. T. Stoddard, Reduced risk of disease during postsecondary dengue virus infections. J. Infect. Dis. 208, 1026–1033 (2013).

28. L. C. Katzelnick, J. V. Zambrana, D. Elizondo, D. Collado, N. Garcia, S. Arguello, J. C. Mercado, T. Miranda, O. Ampie, B. L. Mercado, C. Narvaez, L. Gresh, R. A. Binder, S. Ojeda, N. Sanchez, M. Plazaola, K. Latta, A. Schiller, J. Coloma, F. B. Carrillo, F. Narvaez, M. E. Halloran, A. Gordon, G. Kuan, A. Balmaseda, E. Harris, Dengue and Zika virus infections in children elicit cross-reactive protective and enhancing antibodies that persist long term. Sci. Transl. Med. 13, eabg9478 (2021).

29. Pérez-Guzmán EX, Pantoja P, Serrano-Collazo C, Hassert MA, Ortiz-Rosa A, Rodríguez IV, Giavedoni L, Hodara V, Parodi L, Cruz L, Arana T, White LJ, Martínez MI, Weiskopf D, Brien JD, de Silva A, Pinto AK, Sariol CA, Time elapsed between Zika and dengue virus infections affects antibody and T cell responses. Nat. comm. 10, (2019).

30. N. Marzan-Rivera, C. Serrano-Collazo, L. Cruz, P. Pantoja, A. Ortiz-Rosa, T. Arana, M. I. Martinez, A. G. Burgos, C. Roman, L. B. Mendez, E. Geerling, A. K. Pinto, J. D. Brien, C. A. Sariol, Infection order outweighs the role of CD4(+) T cells in tertiary flavivirus exposure. iScience 25, 104764 (2022).

31. B. M. Valencia, P. C. Sigera, P. Weeratunga, N. Tedla, D. Fernando, S. Rajapakse, A. R. Lloyd, C. Rodrigo, Effect of prior Zika and dengue virus exposure on the severity of a subsequent dengue infection in adults. Sci. Rep. 12, 17225 (2022).

32. P. Bhoomiboonchoo, A. Nisalak, N. Chansatiporn, I. K. Yoon, S. Kalayanarooj, M. Thipayamongkolgul, T. Endy, A. L. Rothman, S. Green, A. Srikiatkhachorn, D. Buddhari, M. P. Mammen, R. V. Gibbons, Sequential dengue virus infections detected in active and passive surveillance programs in Thailand, 1994-2010. BMC Public Health 15, 250 (2015).

33. R. V. Gibbons, S. Kalanarooj, R. G. Jarman, A. Nisalak, D. W. Vaughn, T. P. Endy, M. P. Mammen, Jr., A. Srikiatkhachorn, Analysis of repeat hospital admissions for dengue to estimate the frequency of third or fourth dengue infections resulting in admissions and dengue hemorrhagic fever, and serotype sequences. Am. J. Trop. Med. Hyg. 77, 910–913 (2007).

34. M. Montoya, L. Gresh, J. C. Mercado, K. L. Williams, M. J. Vargas, G. Gutierrez, G. Kuan, A. Gordon, A. Balmaseda, E. Harris, Symptomatic versus inapparent outcome in repeat dengue virus infections is influenced by the time interval between infections and study year. PLoS Negl. Trop. Dis. 7, e2357 (2013).

35. A. Gordon, G. Kuan, J. C. Mercado, L. Gresh, W. Aviles, A. Balmaseda, E. Harris, The Nicaraguan pediatric dengue cohort study: incidence of inapparent and symptomatic dengue virus infections, 2004-2010. PLoS Negl. Trop. Dis. 7, e2462 (2013).

36. T. K. Tsang, S. L. Ghebremariam, L. Gresh, A. Gordon, M. E. Halloran, L. C. Katzelnick, D. P. Rojas, G. Kuan, A. Balmaseda, J. Sugimoto, E. Harris, I. M. Longini, Jr., Y. Yang, Effects of infection history on dengue virus infection and pathogenicity. Nat Commun 10, 1246 (2019).

37. K. M. Soo, B. Khalid, S. M. Ching, H. Y. Chee, Meta-Analysis of Dengue Severity during Infection by Different Dengue Virus Serotypes in Primary and Secondary Infections. PLoS One 11, e0154760 (2016).

38. R. Raut, K. S. Corbett, R. N. Tennekoon, S. Premawansa, A. Wijewickrama, G. Premawansa, P. Mieczkowski, C. Ruckert, G. D. Ebel, A. D. De Silva, A. M. de Silva, Dengue type 1 viruses circulating in humans are highly infectious and poorly neutralized by human antibodies. Proc. Natl. Acad. Sci. U S A 116, 227–232 (2019).

39. K. A. Dowd, C. R. DeMaso, T. C. Pierson, Genotypic Differences in Dengue Virus Neutralization Are Explained by a Single Amino Acid Mutation That Modulates Virus Breathing. mBio 6, e01559–01515 (2015).

40. H. Salje, M. T. Alera, M. N. Chua, T. Hunsawong, D. Ellison, A. Srikiatkhachorn, R. G. Jarman, G. D. Gromowski, I. Rodriguez-Barraquer, S. Cauchemez, D. A. T. Cummings, L. Macareo, I. K. Yoon, S. Fernandez, A. L. Rothman, Evaluation of the extended efficacy of the Dengvaxia vaccine against symptomatic and subclinical dengue infection. Nat. Med. 27, 1395–1400 (2021).

41. L. Lopez, R. E. Paul, V. M. Cao-Lormeau, X. Rodo, Considering waning immunity to better explain dengue dynamics. Epidemics 41, 100630 (2022).

42. K. B. Anderson, R. V. Gibbons, D. A. Cummings, A. Nisalak, S. Green, D. H. Libraty, R. G. Jarman, A. Srikiatkhachorn, M. P. Mammen, B. Darunee, I. K. Yoon, T. P. Endy, A shorter time interval between first and second dengue infections is associated with protection from clinical illness in a school-based cohort in Thailand. J. Infect. Dis. 209, 360–368 (2014).

43. O. Brathwaite Dick, J. L. San Martin, R. H. Montoya, J. del Diego, B. Zambrano, G. H. Dayan, The history of dengue outbreaks in the Americas. Am. J. Trop. Med. Hyg. 87, 584–593 (2012).

44. H. C. Metsky, C. B. Matranga, S. Wohl, S. F. Schaffner, C. A. Freije, S. M. Winnicki, K. West, J. Qu, M. L. Baniecki, A. Gladden-Young, A. E. Lin, C. H. Tomkins-Tinch, S. H. Ye, D. J. Park, C. Y. Luo, K. G. Barnes, R. R. Shah, B. Chak, G. Barbosa-Lima, E. Delatorre, Y. R. Vieira, L. M. Paul, A. L. Tan, C. M. Barcellona, M. C. Porcelli, C. Vasquez, A. C. Cannons, M. R. Cone, K. N. Hogan, E. W. Kopp, J. J. Anzinger, K. F. Garcia, L. A. Parham, R. M. G. Ramirez, M. C. M. Montoya, D. P. Rojas, C. M. Brown, S. Hennigan, B. Sabina, S. Scotland, K. Gangavarapu, N. D. Grubaugh, G. Oliveira, R. Robles-Sikisaka, A. Rambaut, L. Gehrke, S. Smole, M. E. Halloran, L. Villar, S. Mattar, I. Lorenzana, J. Cerbino-Neto, C. Valim, W. Degrave, P. T. Bozza, A. Gnirke, K. G. Andersen, S. Isern, S. F. Michael, F. A. Bozza, T. M. L. Souza, I. Bosch, N. L. Yozwiak, B. L. MacInnis, P. C. Sabeti, Zika virus evolution and spread in the Americas. Nature 546, 411–415 (2017).

45. Q. Zhang, K. Sun, M. Chinazzi, Y. Piontti A. Pastore, N. E. Dean, D. P. Rojas, S. Merler, D. Mistry, P. Poletti, L. Rossi, M. Bray, M. E. Halloran, I. M. Longini, Jr., A. Vespignani, Spread of Zika virus in the Americas. Proc. Natl. Acad. Sci. U S A 114, E4334–E4343 (2017).

46. G. Kuan, A. Gordon, W. Aviles, O. Ortega, S. N. Hammond, D. Elizondo, A. Nunez, J. Coloma, A. Balmaseda, E. Harris, The Nicaraguan pediatric dengue cohort study: study design, methods, use of information technology, and extension to other infectious diseases. Am. J. Epidemiol. 170, 120–129 (2009).

47. A. Gordon, G. Kuan, W. Aviles, N. Sanchez, S. Ojeda, B. Lopez, L. Gresh, A. Balmaseda, E. Harris, The Nicaraguan pediatric influenza cohort study: design, methods, use of technology, and compliance. BMC Infect. Dis. 15, 504 (2015).

48. J. V. Zambrana, F. Bustos Carrillo, R. Burger-Calderon, D. Collado, N. Sanchez, S. Ojeda, J. Carey Monterrey, M. Plazaola, B. Lopez, S. Arguello, D. Elizondo, W. Aviles, J. Coloma, G. Kuan, A. Balmaseda, A. Gordon, E. Harris, Seroprevalence, risk factor, and spatial analyses of Zika virus infection after the 2016 epidemic in Managua, Nicaragua. Proc. Natl. Acad. Sci. U S A 115, 9294–9299 (2018).

49. J. J. Waggoner, L. Gresh, A. Mohamed-Hadley, G. Ballesteros, M. J. Davila, Y. Tellez, M. K. Sahoo, A. Balmaseda, E. Harris, B. A. Pinsky, Single-Reaction Multiplex Reverse Transcription PCR for Detection of Zika, Chikungunya, and Dengue Viruses. Emerg. Infect. Dis. 22, 1295–1297 (2016).

50. A. Balmaseda, E. Sandoval, L. Perez, C. M. Gutierrez, E. Harris, Application of molecular typing techniques in the 1998 dengue epidemic in Nicaragua. Am. J. Trop. Med. Hyg. 61, 893–897 (1999).

51. R. S. Lanciotti, C. H. Calisher, D. J. Gubler, G. J. Chang, A. V. Vorndam, Rapid detection and typing of dengue viruses from clinical samples by using reverse transcriptase-polymerase chain reaction. J Clin Microbiol 30, 545–551 (1992).

52. J. J. Waggoner, G. Ballesteros, L. Gresh, A. Mohamed-Hadley, Y. Tellez, M. K. Sahoo, J. Abeynayake, A. Balmaseda, E. Harris, B. A. Pinsky, Clinical evaluation of a single-reaction real-time RT-PCR for pan-dengue and chikungunya virus detection. J. Clin. Virol. 78, 57–61 (2016).

53. J. J. Waggoner, J. Abeynayake, M. K. Sahoo, L. Gresh, Y. Tellez, K. Gonzalez, G. Ballesteros, A. M. Pierro, P. Gaibani, F. P. Guo, V. Sambri, A. Balmaseda, K. Karunaratne, E. Harris, B. A. Pinsky, Single-reaction, multiplex, real-time rt-PCR for the detection, quantitation, and serotyping of dengue viruses. PLoS Negl. Trop. Dis. 7, e2116 (2013).

54. A. Balmaseda, J. V. Zambrana, D. Collado, N. Garcia, S. Saborio, D. Elizondo, J. C. Mercado, K. Gonzalez, C. Cerpas, A. Nunez, D. Corti, J. J. Waggoner, G. Kuan, R. Burger-Calderon, E. Harris, Comparison of Four Serological Methods and Two Reverse Transcription-PCR Assays for Diagnosis and Surveillance of Zika Virus Infection. J. Clin. Microbiol. 56, 56(3):e01785–17 (2018).

55. A. Balmaseda, M. G. Guzman, S. Hammond, G. Robleto, C. Flores, Y. Tellez, E. Videa, S. Saborio, L. Perez, E. Sandoval, Y. Rodriguez, E. Harris, Diagnosis of dengue virus infection by detection of specific immunoglobulin M (IgM) and IgA antibodies in serum and saliva. Clin. Diagn. Lab. Immunol. 10, 317–322 (2003).

56. A. Gordon, L. Gresh, S. Ojeda, L. C. Katzelnick, N. Sanchez, J. C. Mercado, G. Chowell, B. Lopez, D. Elizondo, J. Coloma, R. Burger-Calderon, G. Kuan, A. Balmaseda, E. Harris, Prior dengue virus infection and risk of Zika: A pediatric cohort in Nicaragua. PLoS Med. 16, e1002726 (2019).

57. Dustin Tingley, Teppei Yamamoto, Kentaro Hirose, Luke Keele, Kosuke Imai, mediation: R Package for Causal Mediation Analysis. J. of Stat. Soft. 59, (2014).

58. R Core Team. R: A Language and Environment for Statistical Computing (2023).

59. Vito M. R. Muggeo, Testing with a nuisance parameter present only under the alternative: a score-based approach with application to segmented modelling. J. of Stat. Comp. and Sim. 86, 3059–3067 (2016).

60. Douglas Bates, Martin Mächler, Ben Bolker, Steve Walker, Fitting Linear Mixed-Effects Models Using lme4. J. of Stat. Soft. 67, 1–48 (2015).

61. D. Alwin, R. Hauser, The Decomposition of Effects in Path Analysis. American Sociological Review, 40, 37–47 (1975).

62. Simon N. Wood, Yannig Goude, Simon Shaw, Generalized Additive Models for Large Data Sets. Journal of the Royal Statistical Society Series C: Applied Statistics 64, 139–155 (2015).

