## Supplemental Material for "Primary exposure to Zika virus increases risk of symptomatic dengue virus infection with serotypes 2, 3, and 4 but not serotype 1"

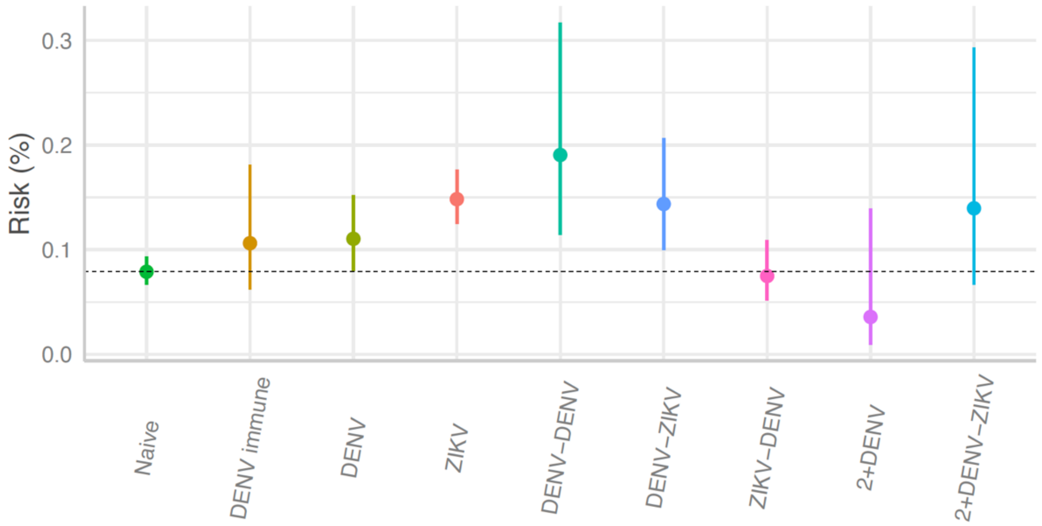


**Fig. S1. Risk of dengue differs by infection history on crude levels.** Crude risk of symptomatic DENV infection by prior DENV and ZIKV infection histories, 2022-2023.

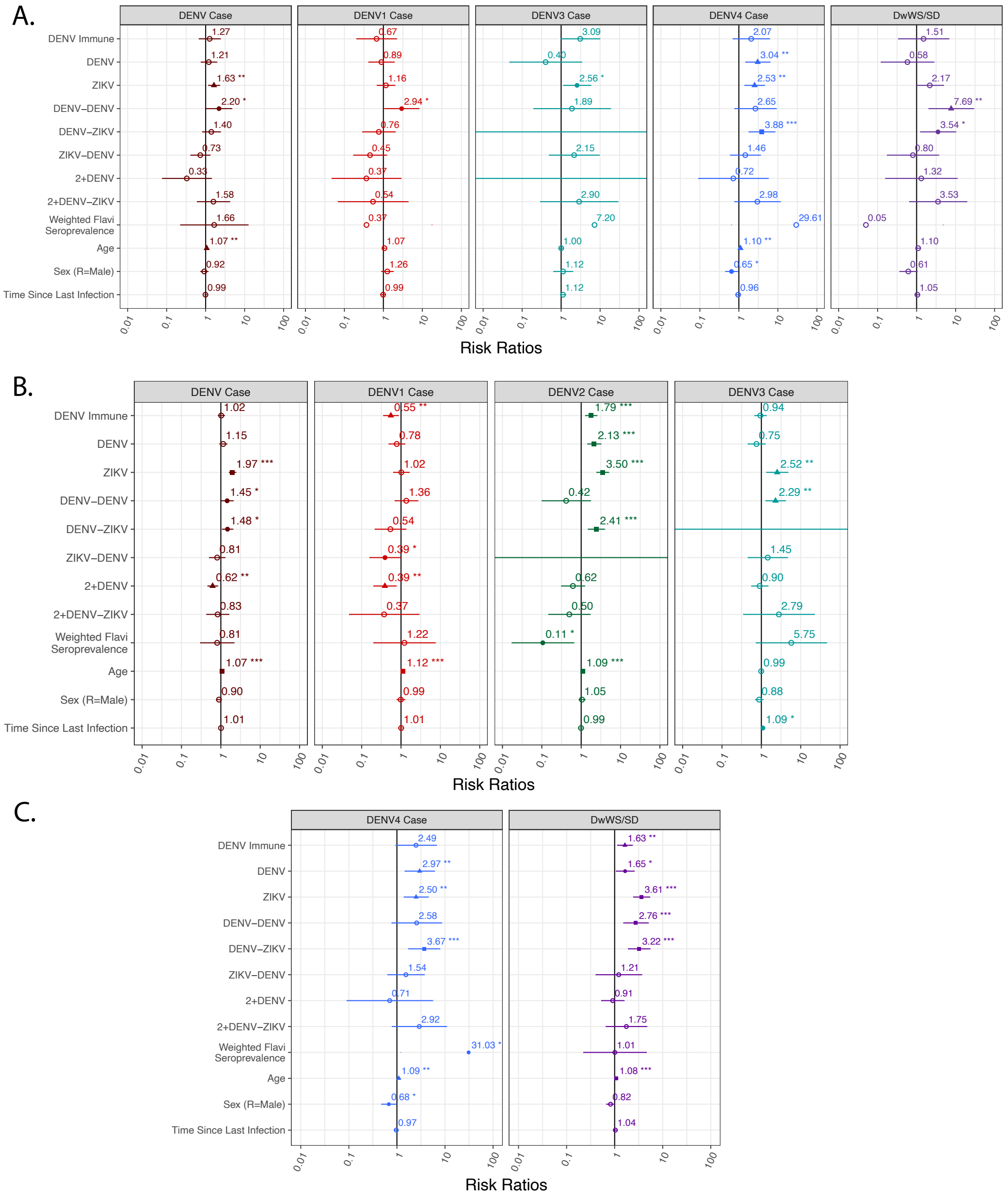


**Fig. S2. Prior infection history confers differential risk of symptomatic DENV1-4 considering baseline spatial risk.** Risks ratios of symptomatic and severe dengue by prior DENV and ZIKV infection histories including age-adjusted flavivirus seroprevalence of the prior year on the neighborhood level as a covariate **(A)** in the 2022-2023 epidemic season **(B-C)** in the entire cohort season (2004-2023).

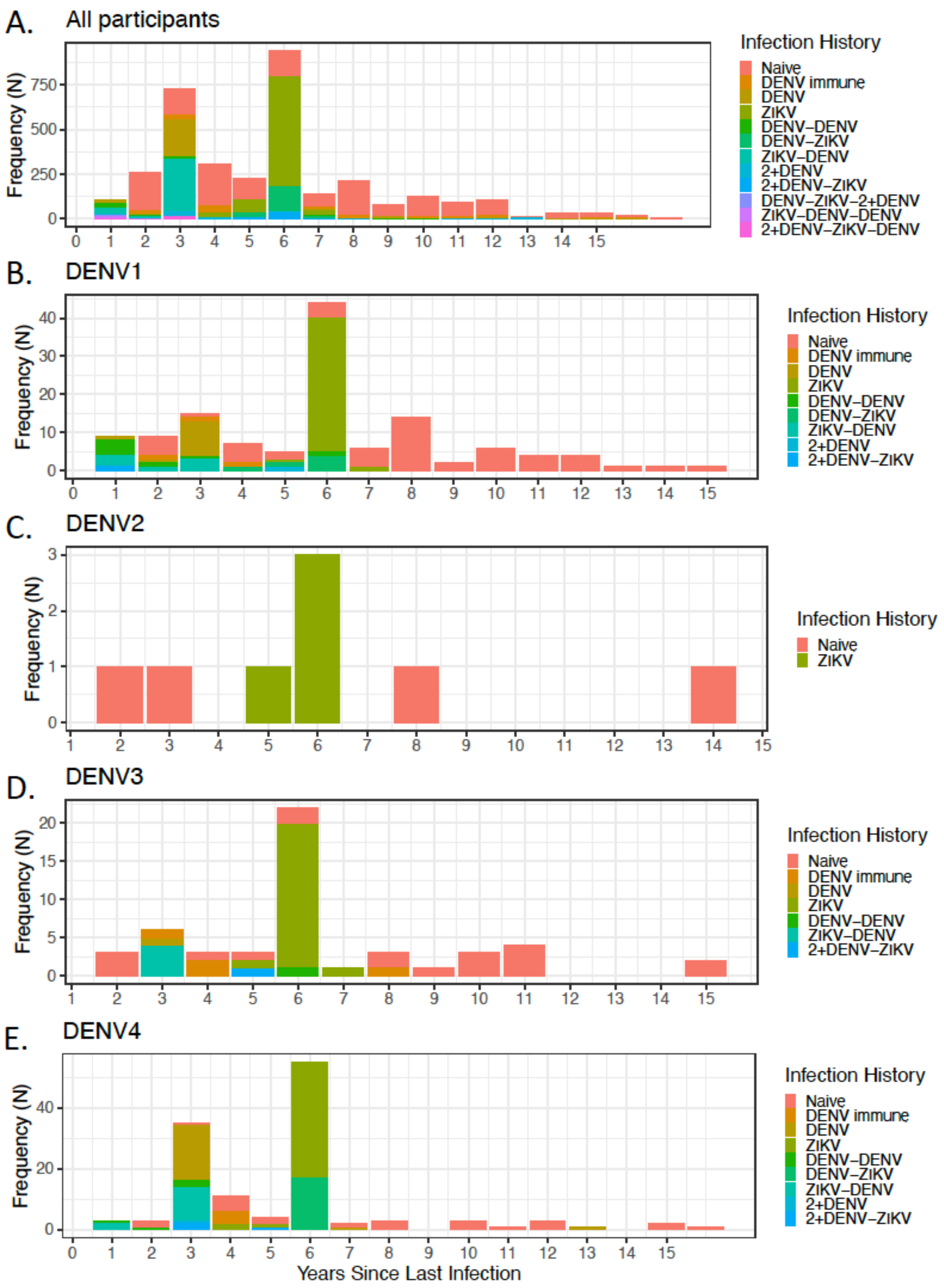


**Fig. S3. Most secondary infections in 2022 had DENV2 and/or ZIKV infections as a primary infection.** Frequency of DENV cases **(A)** and cases due to DENV1 **(B)**, DENV2 **(C)**, DENV3 **(D)**, and DENV4 **(E)** infections in 2022 by years since last infection for each infection history. The flavivirus-naïve group contains years since entered the cohort.

**Table S1. Characteristics of participants in the Pediatric Dengue Cohort Study with documented infection histories, 2004-2023**

| Variable | DENV Cases | | | | | | |
|---|---|---|---|---|---|---|---|
| | Overall, N = 1,418[1] | DENV1, N = 294[1] | DENV2, N = 481[1] | DENV3, N = 310[1] | DENV4, N = 135[1] | Dengue Serology, N = 178[1] | P-value [2] |
| **Age** | 9.5 (6.8, 12.2) | 9.9 (7.4, 12.5) | 9.5 (7.0, 12.1) | 8.8 (6.0, 11.1) | 11.9 (9.8, 14.3) | 8.2 (5.7, 10.7) | <0.001 |
| **Sex** | | | | | | | 0.2 |
| F | 738 (53%) | 149 (51%) | 239 (50%) | 159 (52%) | 78 (58%) | 113 (58%) | |
| M | 667 (47%) | 142 (49%) | 239 (50%) | 148 (48%) | 56 (42%) | 82 (42%) | |
| **DHF/DSS** | 66 (4.7%) | 8 (2.7%) | 30 (6.2%) | 28 (9.0%) | 0 (0%) | 0 (0%) | <0.001 |
| **DwWS/SD** | 383 (27%) | 62 (21%) | 182 (38%) | 103 (33%) | 24 (18%) | 12 (6.1%) | <0.001 |
| **Time Since Last Infection** | 4.54 (3.00, 6.00) | 5.36 (3.00, 8.00) | 3.72 (3.00, 4.00) | 4.70 (3.00, 6.00) | 5.35 (3.00, 6.00) | 4.58 (2.00, 7.00) | <0.001 |
| **Geometric Mean Titer (iELISA)** | 384 (2, 126) | 214 (2, 43) | 514 (2, 197) | 141 (2, 64) | 675 (2, 289) | 491 (2, 39) | <0.001 |
| **Infection History** | | | | | | | |
| Naive | 523 (38%) | 139 (49%) | 115 (24%) | 134 (45%) | 24 (19%) | 111 (58%) | |
| DENV immune | 215 (16%) | 36 (13%) | 86 (18%) | 67 (23%) | 6 (4.7%) | 20 (10%) | |
| DENV | 124 (9.0%) | 29 (10%) | 39 (8.2%) | 21 (7.1%) | 20 (16%) | 15 (7.9%) | |
| ZIKV | 306 (22%) | 39 (14%) | 180 (38%) | 21 (7.1%) | 41 (32%) | 25 (13%) | |
| DENV-DENV | 41 (3.0%) | 13 (4.6%) | 2 (0.4%) | 18 (6.1%) | 4 (3.1%) | 4 (2.1%) | |
| DENV-ZIKV | 71 (5.2%) | 6 (2.1%) | 43 (9.0%) | 0 (0%) | 17 (13%) | 5 (2.6%) | |
| ZIKV-DENV | 26 (1.9%) | 7 (2.5%) | 0 (0%) | 4 (1.3%) | 13 (10%) | 2 (1.0%) | |
| 2+DENV | 60 (4.4%) | 12 (4.3%) | 10 (2.1%) | 31 (10%) | 1 (0.8%) | 6 (3.1%) | |
| 2+DENV-ZIKV | 11 (0.8%) | 1 (0.4%) | 3 (0.6%) | 1 (0.3%) | 3 (2.3%) | 3 (1.6%) | |
| DENV-ZIKV-2+DENV | 0 (0%) | 0 (0%) | 0 (0%) | 0 (0%) | 0 (0%) | 0 (0%) | |
| ZIKV-DENV-DENV | 0 (0%) | 0 (0%) | 0 (0%) | 0 (0%) | 0 (0%) | 0 (0%) | |
| 2+DENV-ZIKV-DENV | 0 (0%) | 0 (0%) | 0 (0%) | 0 (0%) | 0 (0%) | 0 (0%) | |

[1]Mean (IQR); n (%)
[2]Kruskal-Wallis rank sum test; Pearson's Chi-squared test

### Table S2. Estimated risk ratios of a dengue case by prior DENV and ZIKV infection histories, 2022-2023.

| | **DENV Case** | | | **DENV1 Case** | | | **DENV3 Case** | | | **DENV4 Case** | | | **DwWS/SD** | | |
|---|---|---|---|---|---|---|---|---|---|---|---|---|---|---|---|
| *Predictors* | *Risk Ratios* | *CI* | *p* | *Risk Ratios* | *CI* | *p* | *Risk Ratios* | *CI* | *p* | *Risk Ratios* | *CI* | *p* | *Incidence Rate Ratios* | *CI* | *p* |
| (Intercept) | 0.06 | 0.04 – 0.08 | **<0.001** | 0.02 | 0.01 – 0.04 | **<0.001** | 0.01 | 0.00 – 0.02 | **<0.001** | 0.01 | 0.01 – 0.03 | **<0.001** | 0.00 | 0.00 – 0.01 | **<0.001** |
| inf hist DD [DENV immune] | 1.26 | 0.72 – 2.23 | 0.420 | 0.69 | 0.21 – 2.18 | 0.523 | 3.23 | 1.09 – 9.57 | **0.034** | 2.08 | 0.73 – 5.96 | 0.171 | 1.60 | 0.37 – 7.04 | 0.532 |
| inf hist DD [DENV] | 1.25 | 0.82 – 1.89 | 0.296 | 0.91 | 0.44 – 1.88 | 0.801 | 0.38 | 0.05 – 3.00 | 0.359 | 3.25 | 1.65 – 6.41 | **0.001** | 0.60 | 0.13 – 2.81 | 0.515 |
| inf hist DD [ZIKV] | 1.61 | 1.20 – 2.17 | **0.002** | 1.20 | 0.72 – 1.99 | 0.479 | 2.90 | 1.34 – 6.27 | **0.007** | 2.62 | 1.48 – 4.63 | **0.001** | 2.19 | 0.99 – 4.83 | 0.053 |
| inf hist DD [DENV-DENV] | 1.96 | 1.05 – 3.66 | **0.036** | 2.45 | 0.97 – 6.20 | 0.058 | 2.20 | 0.25 – 19.20 | 0.476 | 2.58 | 0.81 – 8.26 | 0.109 | 6.79 | 1.88 – 24.58 | **0.003** |
| inf hist DD [DENV-ZIKV] | 1.43 | 0.90 – 2.29 | 0.131 | 0.76 | 0.30 – 1.93 | 0.567 | 0.00 | 0.00 – Inf | 0.984 | 3.96 | 1.93 – 8.12 | **<0.001** | 3.46 | 1.24 – 9.65 | **0.018** |
| inf hist DD [ZIKV-DENV] | 0.79 | 0.47 – 1.34 | 0.384 | 0.48 | 0.19 – 1.24 | 0.130 | 1.76 | 0.43 – 7.13 | 0.431 | 1.60 | 0.68 – 3.76 | 0.282 | 0.82 | 0.18 – 3.69 | 0.797 |
| inf hist DD [2+DENV] | 0.37 | 0.09 – 1.50 | 0.164 | 0.39 | 0.05 – 2.94 | 0.364 | 0.00 | 0.00 – Inf | 0.991 | 0.78 | 0.10 – 5.98 | 0.815 | 1.37 | 0.17 – 11.06 | 0.768 |
| inf hist DD [2+DENV-ZIKV] | 1.28 | 0.56 – 2.92 | 0.563 | 0.46 | 0.06 – 3.50 | 0.454 | 2.62 | 0.30 – 23.26 | 0.387 | 2.56 | 0.72 – 9.06 | 0.146 | 2.85 | 0.54 – 15.01 | 0.217 |
| inf.hist.DDDENV-ZIKV-2+DENV | 0.00 | 0.00 – Inf | 0.986 | 0.00 | 0.00 – Inf | 0.992 | 0.00 | 0.00 – Inf | 0.998 | 0.00 | 0.00 – Inf | 0.992 | 0.00 | 0.00 – Inf | 0.995 |
| age | 1.04 | 1.00 – 1.08 | **0.029** | 1.05 | 0.98 – 1.12 | 0.150 | 0.98 | 0.88 – 1.09 | 0.687 | 1.07 | 1.01 – 1.14 | **0.023** | 1.07 | 0.98 – 1.18 | 0.141 |
| sex [M] | 0.94 | 0.77 – 1.14 | 0.525 | 1.26 | 0.89 – 1.77 | 0.193 | 1.14 | 0.66 – 1.97 | 0.634 | 0.67 | 0.47 – 0.95 | **0.025** | 0.62 | 0.37 – 1.04 | 0.069 |
| time per imm class | 1.00 | 0.95 – 1.05 | 0.910 | 0.99 | 0.91 – 1.08 | 0.841 | 1.12 | 0.98 – 1.29 | 0.100 | 0.96 | 0.88 – 1.05 | 0.391 | 1.06 | 0.94 – 1.20 | 0.369 |
| Observations | 3411 | | | 3411 | | | 3411 | | | 3411 | | | 3411 | | |
| $R^2$ Nagelkerke | 0.033 | | | 0.022 | | | 0.053 | | | 0.071 | | | 0.087 | | |

### Table S3. Estimated risk ratios of a dengue case by prior DENV and ZIKV infection histories, 2004-2023

| | DENV Case | | | DENV1 Case | | | DENV2 Case | | | DENV3 Case | | | DENV4 Case | | | DwWS/SD | | |
|---|---|---|---|---|---|---|---|---|---|---|---|---|---|---|---|---|---|---|
| *Predictors* | *Incidence Rate Ratios* | *CI* | *p* | *Risk Ratios* | *CI* | *p* | *Risk Ratios* | *CI* | *p* | *Incidence Rate Ratios* | *CI* | *p* | *Incidence Rate Ratios* | *CI* | *p* | *Incidence Rate Ratios* | *CI* | *p* |
| (Intercept) | 0.00 | 0.00 – 0.01 | **<0.001** | 0.00 | 0.00 – 0.00 | **<0.001** | 0.00 | 0.00 – 0.00 | **<0.001** | 0.00 | 0.00 – Inf | 0.996 | 0.00 | 0.00 – 0.00 | **<0.001** | 0.00 | 0.00 – 0.00 | **<0.001** |
| inf hist DD [DENV immune] | 1.04 | 0.87 – 1.25 | 0.649 | 0.57 | 0.38 – 0.86 | **0.008** | 1.84 | 1.33 – 2.55 | **<0.001** | 0.99 | 0.71 – 1.40 | 0.973 | 2.65 | 1.03 – 6.83 | **0.044** | 1.67 | 1.16 – 2.39 | **0.006** |
| inf hist DD [DENV] | 1.17 | 0.94 – 1.45 | 0.160 | 0.80 | 0.51 – 1.27 | 0.346 | 2.09 | 1.44 – 3.05 | **<0.001** | 0.78 | 0.48 – 1.29 | 0.341 | 3.28 | 1.64 – 6.57 | **0.001** | 1.67 | 1.09 – 2.57 | **0.019** |
| inf hist DD [ZIKV] | 1.87 | 1.54 – 2.27 | **<0.001** | 1.04 | 0.66 – 1.63 | 0.856 | 3.62 | 2.60 – 5.04 | **<0.001** | 2.60 | 1.39 – 4.85 | **0.003** | 2.66 | 1.49 – 4.76 | **0.001** | 3.62 | 2.44 – 5.38 | **<0.001** |
| inf hist DD [DENV-DENV] | 1.45 | 1.03 – 2.04 | **0.035** | 1.37 | 0.72 – 2.61 | 0.335 | 0.37 | 0.09 – 1.51 | 0.164 | 2.40 | 1.37 – 4.23 | **0.002** | 2.64 | 0.80 – 8.68 | 0.110 | 2.78 | 1.53 – 5.03 | **0.001** |
| inf hist DD [DENV-ZIKV] | 1.47 | 1.09 – 1.99 | **0.012** | 0.57 | 0.24 – 1.37 | 0.210 | 2.53 | 1.64 – 3.91 | **<0.001** | 0.00 | 0.00 – Inf | 0.997 | 4.05 | 1.93 – 8.49 | **<0.001** | 3.26 | 1.94 – 5.50 | **<0.001** |
| inf hist DD [ZIKV-DENV] | 0.85 | 0.55 – 1.34 | 0.490 | 0.42 | 0.18 – 1.02 | 0.055 | 0.00 | 0.00 – Inf | 0.990 | 1.47 | 0.47 – 4.66 | 0.510 | 1.67 | 0.70 – 4.00 | 0.249 | 1.22 | 0.41 – 3.67 | 0.724 |
| inf hist DD [2+DENV] | 0.64 | 0.48 – 0.86 | **0.003** | 0.41 | 0.22 – 0.79 | **0.008** | 0.55 | 0.28 – 1.09 | 0.086 | 0.96 | 0.60 – 1.56 | 0.883 | 0.76 | 0.10 – 5.86 | 0.790 | 0.93 | 0.54 – 1.60 | 0.785 |
| inf hist DD [2+DENV-ZIKV] | 0.82 | 0.44 – 1.54 | 0.533 | 0.32 | 0.04 – 2.36 | 0.263 | 0.45 | 0.14 – 1.46 | 0.183 | 2.33 | 0.30 – 18.05 | 0.418 | 2.60 | 0.71 – 9.56 | 0.151 | 1.72 | 0.64 – 4.58 | 0.280 |
| inf.hist.DDDENV-ZIKV-2+DENV | 0.00 | 0.00 – Inf | 0.999 | 0.00 | 0.00 – Inf | 1.000 | 0.00 | 0.00 – Inf | 1.000 | 0.00 | 0.00 – Inf | 1.000 | 0.00 | 0.00 – Inf | 1.000 | 0.00 | 0.00 – Inf | 1.000 |
| age | 1.05 | 1.03 – 1.08 | **<0.001** | 1.10 | 1.05 – 1.16 | **<0.001** | 1.09 | 1.05 – 1.12 | **<0.001** | 0.98 | 0.93 – 1.04 | 0.542 | 1.08 | 1.01 – 1.14 | **0.020** | 1.08 | 1.04 – 1.12 | **<0.001** |
| sex [M] | 0.90 | 0.81 – 1.00 | 0.052 | 0.97 | 0.77 – 1.23 | 0.827 | 1.04 | 0.88 – 1.23 | 0.666 | 0.88 | 0.70 – 1.11 | 0.288 | 0.69 | 0.49 – 0.99 | **0.042** | 0.83 | 0.68 – 1.01 | 0.065 |
| time per imm class | 1.01 | 0.98 – 1.04 | 0.421 | 1.01 | 0.96 – 1.08 | 0.627 | 1.00 | 0.95 – 1.06 | 0.968 | 1.09 | 1.01 – 1.18 | **0.020** | 0.97 | 0.88 – 1.06 | 0.482 | 1.04 | 0.98 – 1.10 | 0.242 |
| cohort year [2005] | 3.53 | 2.01 – 6.22 | **<0.001** | 2.58 | 1.16 – 5.73 | **0.020** | 8.03 | 2.44 – 26.41 | **0.001** | 0.94 | 0.00 – Inf | 1.000 | 0.00 | 0.00 – Inf | 1.000 | 1.70 | 0.15 – 18.78 | 0.664 |
| cohort year [2006] | 0.73 | 0.34 – 1.53 | 0.399 | 0.09 | 0.01 – 0.76 | **0.027** | 2.61 | 0.70 – 9.64 | 0.151 | 0.90 | 0.00 – Inf | 1.000 | 0.00 | 0.00 – Inf | 1.000 | 0.80 | 0.05 – 12.79 | 0.873 |
| cohort year [2007] | 3.58 | 2.03 – 6.32 | **<0.001** | 0.09 | 0.01 – 0.74 | **0.025** | 19.11 | 5.96 – 61.28 | **<0.001** | 0.86 | 0.00 – Inf | 1.000 | 0.00 | 0.00 – Inf | 1.000 | 12.26 | 1.61 – 93.44 | **0.016** |
| cohort year [2008] | 1.12 | 0.58 – 2.18 | 0.729 | 0.00 | 0.00 – Inf | 1.000 | 0.56 | 0.09 – 3.40 | 0.532 | 378254773.16 | 0.00 – Inf | 0.997 | 0.00 | 0.00 – Inf | 1.000 | 3.62 | 0.42 – 31.28 | 0.242 |
| cohort year [2009] | 7.80 | 4.54 – 13.39 | **<0.001** | 1.18 | 0.49 – 2.83 | 0.711 | 2.54 | 0.68 – 9.52 | 0.166 | 2448918720.52 | 0.00 – Inf | 0.997 | 0.00 | 0.00 – Inf | 1.000 | 46.10 | 6.31 – 336.96 | **<0.001** |
| cohort year [2010] | 4.67 | 2.67 – 8.16 | **<0.001** | 0.00 | 0.00 – Inf | 1.000 | 4.61 | 1.32 – 16.06 | **0.016** | 1320414757.55 | 0.00 – Inf | 0.997 | 0.00 | 0.00 – Inf | 1.000 | 18.10 | 2.42 – 135.68 | **0.005** |
| cohort year [2011] | 1.46 | 0.76 – 2.79 | 0.255 | 0.31 | 0.09 – 1.06 | 0.063 | 0.00 | 0.00 – Inf | 0.985 | 361967558.36 | 0.00 – Inf | 0.997 | 0.00 | 0.00 – Inf | 1.000 | 3.83 | 0.44 – 33.50 | 0.225 |
| cohort year [2012] | 4.28 | 2.43 – 7.54 | **<0.001** | 5.05 | 2.31 – 11.04 | **<0.001** | 0.28 | 0.03 – 2.73 | 0.273 | 37368050.46 | 0.00 – Inf | 0.997 | 0.00 | 0.00 – Inf | 1.000 | 14.08 | 1.84 – 107.44 | **0.011** |
| cohort year [2013] | 1.75 | 0.94 – 3.27 | 0.080 | 1.28 | 0.53 – 3.08 | 0.582 | 1.79 | 0.43 – 7.34 | 0.421 | 77476627.39 | 0.00 – Inf | 0.997 | 1.04 | 0.06 – 18.11 | 0.979 | 8.53 | 1.08 – 67.53 | **0.042** |
| cohort year [2014] | 0.58 | 0.26 – 1.28 | 0.174 | 0.00 | 0.00 – Inf | 1.000 | 0.30 | 0.03 – 2.99 | 0.307 | 58115982.30 | 0.00 – Inf | 0.997 | 0.00 | 0.00 – Inf | 1.000 | 0.00 | 0.00 – Inf | 0.999 |
| cohort year [2015] | 1.51 | 0.80 – 2.86 | 0.202 | 0.00 | 0.00 – Inf | 1.000 | 7.23 | 2.11 – 24.78 | **0.002** | 0.69 | 0.00 – Inf | 1.000 | 0.00 | 0.00 – Inf | 1.000 | 3.48 | 0.39 – 30.74 | 0.262 |
| cohort year [2016] | 1.60 | 0.85 – 2.99 | 0.142 | 0.00 | 0.00 – Inf | 0.999 | 2.48 | 0.65 – 9.45 | 0.184 | 0.69 | 0.00 – Inf | 1.000 | 0.00 | 0.00 – Inf | 1.000 | 0.00 | 0.00 – Inf | 1.000 |
| cohort year [2017] | 0.15 | 0.05 – 0.44 | **0.001** | 0.00 | 0.00 – Inf | 0.999 | 0.00 | 0.00 – Inf | 0.983 | 0.61 | 0.00 – Inf | 1.000 | 0.00 | 0.00 – Inf | 1.000 | 0.00 | 0.00 – Inf | 0.999 |
| cohort year [2018] | 0.36 | 0.16 – 0.81 | **0.014** | 0.00 | 0.00 – Inf | 0.999 | 0.44 | 0.09 – 2.26 | 0.325 | 0.57 | 0.00 – Inf | 1.000 | 0.00 | 0.00 – Inf | 1.000 | 0.00 | 0.00 – Inf | 0.999 |
| cohort year [2019] | 11.52 | 6.67 – 19.89 | **<0.001** | 0.00 | 0.00 – Inf | 1.000 | 48.67 | 14.75 – 160.55 | **<0.001** | 0.55 | 0.00 – Inf | 1.000 | 0.00 | 0.00 – Inf | 1.000 | 56.26 | 7.54 – 419.97 | **<0.001** |
| cohort year [2020] | 0.19 | 0.07 – 0.53 | **0.002** | 0.00 | 0.00 – Inf | 0.999 | 0.38 | 0.06 – 2.35 | 0.297 | 0.58 | 0.00 – Inf | 1.000 | 0.00 | 0.00 – Inf | 1.000 | 0.00 | 0.00 – Inf | 0.999 |
| cohort year [2021] | 0.12 | 0.03 – 0.41 | **0.001** | 0.10 | 0.02 – 0.50 | **0.005** | 0.00 | 0.00 – Inf | 0.983 | 0.56 | 0.00 – Inf | 1.000 | 0.00 | 0.00 – Inf | 1.000 | 0.00 | 0.00 – Inf | 1.000 |
| cohort year [2022] | 13.06 | 7.49 – 22.77 | **<0.001** | 6.76 | 3.00 – 15.27 | **<0.001** | 1.60 | 0.40 – 6.42 | 0.508 | 705881112.68 | 0.00 – Inf | 0.997 | 97.75 | 11.66 – 819.56 | **<0.001** | 24.68 | 3.22 – 189.12 | **0.002** |
| **Random Effects** | | | | | | | | | | | | | | | | | | |
| $\sigma^2$ | 5.05 | | | NA | | | NA | | | 6.76 | | | 7.59 | | | 0.00 | | |
| $\tau_{00}$ | 0.00 $_{case.ids}$ | | | 0.00 $_{case.ids}$ | | | 1.00 $_{case.ids}$ | | | 0.00 $_{case.ids}$ | | | 0.00 $_{case.ids}$ | | | 0.00 $_{case.ids}$ | | |
| ICC | 0.00 | | | | | | | | | | | | 0.00 | | | 1.00 | | |
| N | 9165 $_{case.ids}$ | | | 9165 $_{case.ids}$ | | | 9165 $_{case.ids}$ | | | 9165 $_{case.ids}$ | | | 9165 $_{case.ids}$ | | | 9165 $_{case.ids}$ | | |
| Observations | 66842 | | | 67438 | | | 67438 | | | 67438 | | | 67438 | | | 67438 | | |
| Marginal $R^2$ / Conditional $R^2$ | 0.303 / 0.303 | | | NA | | | NA | | | 0.943 / NA | | | 0.909 / 0.909 | | | 1.000 / 1.000 | | |

### Table S4. Estimated risk of a dengue case by pre-existing flavivirus immunity, 2004-2023.

| | DENV Case | | | DENV1 Case | | | DENV2 Case | | | DENV3 Case | | | DENV4 Case | | | DwWS/SD | | | DHF/DSS | | |
|---|---|---|---|---|---|---|---|---|---|---|---|---|---|---|---|---|---|---|---|---|---|
| *Predictors* | *Risk Ratios* | *CI* | *p* | *Risk Ratios* | *CI* | *p* | *Risk Ratios* | *CI* | *p* | *Risk Ratios* | *CI* | *p* | *Incidence Rate Ratios* | *CI* | *p* | *Risk Ratios* | *CI* | *p* | *Risk Ratios* | *CI* | *p* |
| (Intercept) | 0.00 | 0.00 – 0.01 | **<0.001** | 0.00 | 0.00 – 0.00 | **<0.001** | 0.00 | 0.00 – 0.00 | **<0.001** | 0.00 | 0.00 – Inf | 0.994 | 0.00 | 0.00 – 0.00 | **<0.001** | 0.00 | 0.00 – 0.00 | **<0.001** | 0.00 | 0.00 – Inf | 0.994 |
| naive TF [FALSE] | 1.28 | 1.12 – 1.47 | **<0.001** | 0.72 | 0.53 – 0.98 | **0.036** | 2.25 | 1.75 – 2.90 | **<0.001** | 1.09 | 0.82 – 1.46 | 0.551 | 2.96 | 1.73 – 5.09 | **<0.001** | 2.16 | 1.63 – 2.87 | **<0.001** | 4.57 | 2.06 – 10.12 | **<0.001** |
| age | 1.04 | 1.02 – 1.06 | **<0.001** | 1.09 | 1.04 – 1.15 | **<0.001** | 1.06 | 1.03 – 1.09 | **<0.001** | 0.99 | 0.94 – 1.04 | 0.644 | 1.06 | 1.00 – 1.12 | **0.038** | 1.07 | 1.03 – 1.11 | **<0.001** | 1.04 | 0.94 – 1.14 | 0.432 |
| sex [M] | 0.89 | 0.80 – 0.99 | **0.026** | 0.96 | 0.76 – 1.21 | 0.749 | 1.00 | 0.84 – 1.19 | 0.964 | 0.88 | 0.71 – 1.11 | 0.287 | 0.68 | 0.48 – 0.97 | **0.033** | 0.81 | 0.67 – 0.99 | **0.040** | 1.13 | 0.69 – 1.87 | 0.620 |
| time per imm class | 1.01 | 0.99 – 1.04 | 0.271 | 1.02 | 0.97 – 1.07 | 0.479 | 0.99 | 0.94 – 1.03 | 0.598 | 1.08 | 1.01 – 1.15 | **0.023** | 1.01 | 0.93 – 1.08 | 0.877 | 1.02 | 0.97 – 1.07 | 0.441 | 1.04 | 0.92 – 1.19 | 0.517 |
| cohort year [2005] | 3.50 | 1.99 – 6.15 | **<0.001** | 2.59 | 1.17 – 5.74 | **0.019** | 7.86 | 2.39 – 25.82 | **0.001** | 0.01 | 0.00 – Inf | 1.000 | 0.00 | 0.00 – Inf | 1.000 | 1.69 | 0.15 – 18.66 | 0.667 | 0.95 | 0.00 – Inf | 1.000 |
| cohort year [2006] | 0.71 | 0.34 – 1.49 | 0.363 | 0.10 | 0.01 – 0.76 | **0.027** | 2.48 | 0.67 – 9.16 | 0.172 | 0.01 | 0.00 – Inf | 1.000 | 0.00 | 0.00 – Inf | 1.000 | 0.78 | 0.05 – 12.52 | 0.863 | 9039689.63 | 0.00 – Inf | 0.996 |
| cohort year [2007] | 3.53 | 2.01 – 6.20 | **<0.001** | 0.09 | 0.01 – 0.75 | **0.026** | 17.68 | 5.55 – 56.35 | **<0.001** | 0.00 | 0.00 – Inf | 1.000 | 0.00 | 0.00 – Inf | 1.000 | 12.27 | 1.62 – 93.02 | **0.015** | 76819368.50 | 0.00 – Inf | 0.996 |
| cohort year [2008] | 1.12 | 0.58 – 2.15 | 0.745 | 0.00 | 0.00 – Inf | 0.995 | 0.54 | 0.09 – 3.22 | 0.496 | 158878004.82 | 0.00 – Inf | 0.996 | 0.00 | 0.00 – Inf | 1.000 | 3.65 | 0.43 – 31.33 | 0.238 | 34717089.91 | 0.00 – Inf | 0.996 |
| cohort year [2009] | 7.76 | 4.56 – 13.20 | **<0.001** | 1.25 | 0.53 – 2.94 | 0.615 | 2.38 | 0.64 – 8.82 | 0.195 | 1052570849.20 | 0.00 – Inf | 0.995 | 0.00 | 0.00 – Inf | 1.000 | 47.00 | 6.50 – 340.01 | **<0.001** | 135079982.71 | 0.00 – Inf | 0.996 |
| cohort year [2010] | 4.63 | 2.68 – 8.01 | **<0.001** | 0.00 | 0.00 – Inf | 0.995 | 4.28 | 1.25 – 14.69 | **0.021** | 567938593.01 | 0.00 – Inf | 0.995 | 0.00 | 0.00 – Inf | 1.000 | 18.43 | 2.49 – 136.26 | **0.004** | 70898226.67 | 0.00 – Inf | 0.996 |
| cohort year [2012] | 4.42 | 2.54 – 7.67 | **<0.001** | 5.58 | 2.64 – 11.81 | **<0.001** | 0.28 | 0.03 – 2.71 | 0.273 | 16314165.16 | 0.00 – Inf | 0.996 | 0.00 | 0.00 – Inf | 1.000 | 15.10 | 2.01 – 113.11 | **0.008** | 35646078.18 | 0.00 – Inf | 0.996 |
| cohort year [2013] | 1.84 | 1.00 – 3.40 | 0.050 | 1.46 | 0.63 – 3.40 | 0.383 | 1.84 | 0.46 – 7.43 | 0.389 | 34169082.89 | 0.00 – Inf | 0.996 | 0.89 | 0.05 – 14.39 | 0.933 | 9.42 | 1.21 – 72.99 | **0.032** | 9898451.86 | 0.00 – Inf | 0.996 |
| cohort year [2014] | 0.62 | 0.28 – 1.36 | 0.231 | 0.00 | 0.00 – Inf | 0.996 | 0.33 | 0.03 – 3.18 | 0.336 | 26031392.05 | 0.00 – Inf | 0.996 | 0.00 | 0.00 – Inf | 1.000 | 0.00 | 0.00 – Inf | 0.994 | 0.94 | 0.00 – Inf | 1.000 |
| cohort year [2015] | 1.66 | 0.89 – 3.08 | 0.111 | 0.00 | 0.00 – Inf | 0.995 | 8.14 | 2.43 – 27.25 | **0.001** | 0.00 | 0.00 – Inf | 1.000 | 0.00 | 0.00 – Inf | 1.000 | 4.09 | 0.47 – 35.35 | 0.200 | 10887801.45 | 0.00 – Inf | 0.996 |
| cohort year [2016] | 1.78 | 0.97 – 3.29 | 0.064 | 0.00 | 0.00 – Inf | 0.995 | 2.89 | 0.78 – 10.77 | 0.113 | 0.00 | 0.00 – Inf | 1.000 | 0.00 | 0.00 – Inf | 1.000 | 0.00 | 0.00 – Inf | 0.993 | 1.13 | 0.00 – Inf | 1.000 |
| cohort year [2018] | 0.50 | 0.22 – 1.11 | 0.087 | 0.00 | 0.00 – Inf | 0.995 | 0.75 | 0.15 – 3.74 | 0.730 | 0.01 | 0.00 – Inf | 1.000 | 0.00 | 0.00 – Inf | 1.000 | 0.00 | 0.00 – Inf | 0.993 | 0.84 | 0.00 – Inf | 1.000 |
| cohort year [2019] | 16.04 | 9.54 – 26.96 | **<0.001** | 0.00 | 0.00 – Inf | 0.995 | 69.78 | 22.28 – 218.53 | **<0.001** | 0.02 | 0.00 – Inf | 1.000 | 0.00 | 0.00 – Inf | 1.000 | 99.96 | 13.92 – 717.64 | **<0.001** | 117811787.88 | 0.00 – Inf | 0.996 |
| cohort year [2020] | 0.24 | 0.09 – 0.66 | **0.006** | 0.00 | 0.00 – Inf | 0.995 | 0.46 | 0.08 – 2.73 | 0.390 | 0.01 | 0.00 – Inf | 1.000 | 0.00 | 0.00 – Inf | 1.000 | 0.00 | 0.00 – Inf | 0.993 | 0.77 | 0.00 – Inf | 1.000 |
| cohort year [2022] | 16.49 | 9.75 – 27.88 | **<0.001** | 8.28 | 3.90 – 17.55 | **<0.001** | 1.99 | 0.52 – 7.59 | 0.313 | 409982817.97 | 0.00 – Inf | 0.995 | 88.54 | 12.01 – 652.74 | **<0.001** | 38.61 | 5.29 – 281.70 | **<0.001** | 23511662.88 | 0.00 – Inf | 0.996 |
| **Random Effects** | | | | | | | | | | | | | | | | | | | | | |
| $\sigma^2$ | NA | | | NA | | | NA | | | NA | | | 7.59 | | | NA | | | NA | | |
| $\tau_{00}$ | 0.00 $_{case.ids}$ | | | 0.00 $_{case.ids}$ | | | 0.00 $_{case.ids}$ | | | 0.00 $_{case.ids}$ | | | 0.00 $_{case.ids}$ | | | 0.00 $_{case.ids}$ | | | 0.00 $_{case.ids}$ | | |
| N | 9165 $_{case.ids}$ | | | 9165 $_{case.ids}$ | | | 9165 $_{case.ids}$ | | | 9165 $_{case.ids}$ | | | 9165 $_{case.ids}$ | | | 9165 $_{case.ids}$ | | | 9165 $_{case.ids}$ | | |
| Observations | 66842 | | | 67438 | | | 67438 | | | 67438 | | | 67438 | | | 67438 | | | 67438 | | |
| Marginal $R^2$ / Conditional $R^2$ | NA | | | NA | | | NA | | | NA | | | 1.000 / NA | | | NA | | | NA | | |

#### Table S5. Estimated risk of symptomatic DENV infection by pre-existing DENV iELISA titers, 2022-2023.

| | DENV Case | | | DENV1 Case | | | DENV3 Case | | | DENV4 Case | | | DwWS/SD | | |
|---|---|---|---|---|---|---|---|---|---|---|---|---|---|---|---|
| *Predictors* | *Risk Ratios* | *CI* | *p* | *Risk Ratios* | *CI* | *p* | *Risk Ratios* | *CI* | *p* | *Risk Ratios* | *CI* | *p* | *Risk Ratios* | *CI* | *p* |
| (Intercept) | 0.06 | 0.04 – 0.08 | **<0.001** | 0.02 | 0.01 – 0.04 | **<0.001** | 0.01 | 0.00 – 0.02 | **<0.001** | 0.01 | 0.01 – 0.02 | **<0.001** | 0.00 | 0.00 – 0.01 | **<0.001** |
| gm titers cat [<21] | 1.62 | 1.21 – 2.17 | **0.001** | 1.10 | 0.65 – 1.82 | 0.714 | 2.91 | 1.39 – 6.09 | **0.005** | 2.76 | 1.55 – 4.96 | **0.001** | 2.16 | 1.01 – 4.61 | **0.047** |
| gm.titers.cat21-80 | 1.64 | 1.12 – 2.34 | **0.009** | 1.27 | 0.66 – 2.32 | 0.459 | 1.90 | 0.69 – 5.26 | 0.215 | 3.59 | 1.84 – 6.89 | **<0.001** | 1.67 | 0.59 – 4.23 | 0.305 |
| gm.titers.cat81-320 | 1.39 | 0.92 – 2.06 | 0.106 | 0.79 | 0.35 – 1.61 | 0.542 | 0.68 | 0.15 – 3.21 | 0.629 | 4.22 | 2.18 – 8.11 | **<0.001** | 1.80 | 0.62 – 4.71 | 0.254 |
| gm.titers.cat321-1280 | 1.10 | 0.69 – 1.70 | 0.670 | 0.72 | 0.31 – 1.54 | 0.430 | 1.08 | 0.28 – 4.18 | 0.906 | 3.07 | 1.46 – 6.28 | **0.003** | 2.26 | 0.82 – 5.79 | 0.102 |
| gm titers cat [>1280] | 0.68 | 0.41 – 1.11 | 0.134 | 0.38 | 0.14 – 0.91 | **0.041** | 0.00 | 0.00 – Inf | 0.984 | 2.08 | 0.94 – 4.49 | 0.067 | 0.69 | 0.17 – 2.32 | 0.572 |
| age | 1.04 | 1.00 – 1.07 | **0.040** | 1.05 | 0.99 – 1.12 | 0.113 | 1.00 | 0.90 – 1.10 | 0.961 | 1.05 | 0.99 – 1.11 | 0.084 | 1.10 | 1.01 – 1.19 | **0.032** |
| sex [M] | 0.91 | 0.74 – 1.10 | 0.326 | 1.21 | 0.86 – 1.71 | 0.272 | 1.10 | 0.64 – 1.91 | 0.719 | 0.66 | 0.46 – 0.93 | **0.020** | 0.62 | 0.37 – 1.01 | 0.060 |
| time per imm class | 1.01 | 0.97 – 1.06 | 0.614 | 0.99 | 0.92 – 1.07 | 0.832 | 1.09 | 0.97 – 1.23 | 0.165 | 1.01 | 0.93 – 1.08 | 0.891 | 1.05 | 0.94 – 1.16 | 0.378 |
| Observations | 3499 | | | 3499 | | | 3499 | | | 3499 | | | 3499 | | |
| $R^2$ Nagelkerke | 0.027 | | | 0.015 | | | 0.056 | | | 0.059 | | | 0.048 | | |

### Table S6. Estimated risk of symptomatic DENV infection by pre-existing DENV iELISA titers, 2004-2023.

| | **DENV Case** | | | **DENV1 Case** | | | **DENV2 Case** | | | **DENV3 Case** | | | **DENV4 Case** | | | **DwWS/SD** | | |
|---|---|---|---|---|---|---|---|---|---|---|---|---|---|---|---|---|---|---|
| *Predictors* | *Risk Ratios* | *CI* | *p* | *Risk Ratios* | *CI* | *p* | *Incidence Rate Ratios* | *CI* | *p* | *Risk Ratios* | *CI* | *p* | *Incidence Rate Ratios* | *CI* | *p* | *Risk Ratios* | *CI* | *p* |
| (Intercept) | 0.00 | 0.00 – 0.01 | **<0.001** | 0.00 | 0.00 – 0.00 | **<0.001** | 0.00 | 0.00 – 0.00 | **<0.001** | 0.00 | 0.00 – Inf | 0.994 | 0.00 | 0.00 – 0.00 | **<0.001** | 0.00 | 0.00 – 0.00 | **<0.001** |
| gm titers cat [<21] | 1.37 | 1.15 – 1.63 | **<0.001** | 0.89 | 0.61 – 1.31 | 0.560 | 1.71 | 1.19 – 2.45 | **0.003** | 1.67 | 1.16 – 2.42 | **0.006** | 2.90 | 1.61 – 5.22 | **<0.001** | 1.83 | 1.27 – 2.64 | **0.001** |
| gm.titers.cat21-80 | 1.62 | 1.37 – 1.92 | **<0.001** | 0.95 | 0.64 – 1.40 | 0.796 | 3.32 | 2.47 – 4.48 | **<0.001** | 1.22 | 0.84 – 1.77 | 0.286 | 3.59 | 1.83 – 7.03 | **<0.001** | 2.97 | 2.15 – 4.10 | **<0.001** |
| gm.titers.cat81-320 | 1.33 | 1.12 – 1.58 | **0.001** | 0.62 | 0.41 – 0.95 | **0.028** | 2.71 | 2.01 – 3.67 | **<0.001** | 0.94 | 0.64 – 1.38 | 0.767 | 4.45 | 2.29 – 8.65 | **<0.001** | 2.34 | 1.67 – 3.27 | **<0.001** |
| gm.titers.cat321-1280 | 0.96 | 0.77 – 1.20 | 0.721 | 0.35 | 0.19 – 0.63 | **<0.001** | 2.24 | 1.57 – 3.19 | **<0.001** | 0.51 | 0.29 – 0.90 | **0.019** | 3.14 | 1.50 – 6.58 | **0.002** | 1.68 | 1.12 – 2.53 | **0.013** |
| gm titers cat [>1280] | 0.64 | 0.48 – 0.84 | **0.002** | 0.25 | 0.12 – 0.52 | **<0.001** | 0.99 | 0.59 – 1.66 | 0.958 | 0.44 | 0.20 – 0.94 | **0.034** | 2.13 | 0.96 – 4.71 | 0.062 | 1.02 | 0.59 – 1.76 | 0.932 |
| age | 1.06 | 1.04 – 1.08 | **<0.001** | 1.11 | 1.06 – 1.17 | **<0.001** | 1.08 | 1.04 – 1.11 | **<0.001** | 1.01 | 0.96 – 1.06 | 0.672 | 1.05 | 0.99 – 1.11 | 0.078 | 1.08 | 1.05 – 1.12 | **<0.001** |
| sex [M] | 0.88 | 0.79 – 0.98 | **0.015** | 0.94 | 0.75 – 1.19 | 0.620 | 1.01 | 0.84 – 1.21 | 0.951 | 0.88 | 0.70 – 1.10 | 0.262 | 0.68 | 0.48 – 0.97 | **0.034** | 0.81 | 0.66 – 0.99 | **0.039** |
| time per imm class | 1.00 | 0.98 – 1.03 | 0.759 | 1.00 | 0.94 – 1.05 | 0.892 | 0.98 | 0.94 – 1.03 | 0.483 | 1.06 | 1.00 – 1.13 | 0.071 | 1.01 | 0.94 – 1.09 | 0.757 | 1.01 | 0.96 – 1.06 | 0.773 |
| cohort year [2005] | 3.50 | 1.99 – 6.14 | **<0.001** | 2.61 | 1.18 – 5.77 | **0.018** | 7.80 | 2.37 – 25.74 | **0.001** | 0.00 | 0.00 – Inf | 1.000 | 0.00 | 0.00 – Inf | 0.999 | 1.69 | 0.15 – 18.65 | 0.668 |
| cohort year [2006] | 0.70 | 0.34 – 1.48 | 0.354 | 0.10 | 0.01 – 0.76 | **0.027** | 2.47 | 0.67 – 9.13 | 0.176 | 0.00 | 0.00 – Inf | 1.000 | 0.00 | 0.00 – Inf | 1.000 | 0.78 | 0.05 – 12.48 | 0.861 |
| cohort year [2007] | 3.40 | 1.94 – 5.96 | **<0.001** | 0.09 | 0.01 – 0.72 | **0.023** | 17.01 | 5.31 – 54.45 | **<0.001** | 0.00 | 0.00 – Inf | 1.000 | 0.00 | 0.00 – Inf | 1.000 | 11.85 | 1.56 – 89.82 | **0.017** |
| cohort year [2008] | 1.05 | 0.55 – 2.03 | 0.876 | 0.00 | 0.00 – Inf | 0.995 | 0.51 | 0.09 – 3.09 | 0.467 | 136901815.68 | 0.00 – Inf | 0.996 | 0.00 | 0.00 – Inf | 0.999 | 3.49 | 0.41 – 29.96 | 0.255 |
| cohort year [2009] | 7.28 | 4.28 – 12.38 | **<0.001** | 1.16 | 0.49 – 2.74 | 0.731 | 2.19 | 0.59 – 8.16 | 0.242 | 915708882.92 | 0.00 – Inf | 0.995 | 0.00 | 0.00 – Inf | 1.000 | 43.85 | 6.06 – 317.38 | **<0.001** |
| cohort year [2010] | 4.36 | 2.52 – 7.55 | **<0.001** | 0.00 | 0.00 – Inf | 0.995 | 4.00 | 1.16 – 13.81 | **0.028** | 486842043.10 | 0.00 – Inf | 0.995 | 0.00 | 0.00 – Inf | 0.999 | 17.43 | 2.36 – 128.92 | **0.005** |
| cohort year [2011] | 1.36 | 0.72 – 2.57 | 0.346 | 0.31 | 0.09 – 1.03 | 0.055 | 0.00 | 0.00 – Inf | 0.999 | 128772175.51 | 0.00 – Inf | 0.996 | 0.00 | 0.00 – Inf | 1.000 | 3.70 | 0.43 – 31.86 | 0.234 |
| cohort year [2012] | 4.04 | 2.33 – 7.02 | **<0.001** | 5.02 | 2.37 – 10.65 | **<0.001** | 0.25 | 0.03 – 2.47 | 0.238 | 13462059.59 | 0.00 – Inf | 0.996 | 0.00 | 0.00 – Inf | 0.999 | 13.74 | 1.83 – 103.04 | **0.011** |
| cohort year [2013] | 1.71 | 0.93 – 3.16 | 0.085 | 1.34 | 0.57 – 3.13 | 0.498 | 1.69 | 0.42 – 6.85 | 0.461 | 28828392.88 | 0.00 – Inf | 0.996 | 0.87 | 0.05 – 14.15 | 0.923 | 8.69 | 1.12 – 67.38 | **0.039** |
| cohort year [2014] | 0.58 | 0.26 – 1.27 | 0.172 | 0.00 | 0.00 – Inf | 0.996 | 0.30 | 0.03 – 2.91 | 0.299 | 22172025.15 | 0.00 – Inf | 0.996 | 0.00 | 0.00 – Inf | 1.000 | 0.00 | 0.00 – Inf | 0.999 |
| cohort year [2015] | 1.56 | 0.84 – 2.91 | 0.160 | 0.00 | 0.00 – Inf | 0.995 | 7.59 | 2.26 – 25.53 | **0.001** | 0.00 | 0.00 – Inf | 1.000 | 0.00 | 0.00 – Inf | 1.000 | 3.84 | 0.44 – 33.17 | 0.221 |
| cohort year [2016] | 1.70 | 0.92 – 3.14 | 0.088 | 0.00 | 0.00 – Inf | 0.995 | 2.83 | 0.76 – 10.61 | 0.122 | 0.00 | 0.00 – Inf | 1.000 | 0.00 | 0.00 – Inf | 0.999 | 0.00 | 0.00 – Inf | 0.999 |
| cohort year [2017] | 0.19 | 0.06 – 0.56 | **0.003** | 0.00 | 0.00 – Inf | 0.995 | 0.00 | 0.00 – Inf | 0.999 | 0.00 | 0.00 – Inf | 1.000 | 0.00 | 0.00 – Inf | 0.999 | 0.00 | 0.00 – Inf | 0.999 |
| cohort year [2018] | 0.46 | 0.20 – 1.02 | 0.055 | 0.00 | 0.00 – Inf | 0.995 | 0.75 | 0.15 – 3.76 | 0.732 | 0.00 | 0.00 – Inf | 1.000 | 0.00 | 0.00 – Inf | 0.999 | 0.00 | 0.00 – Inf | 0.999 |
| cohort year [2019] | 14.44 | 8.58 – 24.31 | **<0.001** | 0.00 | 0.00 – Inf | 0.995 | 64.24 | 20.40 – 202.35 | **<0.001** | 0.02 | 0.00 – Inf | 1.000 | 0.00 | 0.00 – Inf | 0.999 | 91.08 | 12.67 – 654.66 | **<0.001** |
| cohort year [2020] | 0.23 | 0.08 – 0.64 | **0.005** | 0.00 | 0.00 – Inf | 0.995 | 0.47 | 0.08 – 2.85 | 0.414 | 0.00 | 0.00 – Inf | 1.000 | 0.00 | 0.00 – Inf | 1.000 | 0.00 | 0.00 – Inf | 1.000 |
| cohort year [2021] | 0.14 | 0.04 – 0.49 | **0.002** | 0.11 | 0.02 – 0.55 | **0.007** | 0.00 | 0.00 – Inf | 0.999 | 0.02 | 0.00 – Inf | 1.000 | 0.00 | 0.00 – Inf | 1.000 | 0.00 | 0.00 – Inf | 0.999 |
| cohort year [2022] | 15.37 | 9.07 – 26.04 | **<0.001** | 7.42 | 3.48 – 15.81 | **<0.001** | 1.97 | 0.51 – 7.57 | 0.322 | 317888094.00 | 0.00 – Inf | 0.995 | 93.98 | 12.72 – 694.26 | **<0.001** | 37.86 | 5.18 – 276.75 | **<0.001** |
| **Random Effects** | | | | | | | | | | | | | | | | | | |
| $\sigma^2$ | NA | | | NA | | | 6.32 | | | NA | | | 7.59 | | | NA | | |
| $\tau_{00}$ | $0.00_{\text{case.ids}}$ | | | $0.00_{\text{case.ids}}$ | | | $0.00_{\text{case.ids}}$ | | | $0.00_{\text{case.ids}}$ | | | $0.00_{\text{case.ids}}$ | | | $0.00_{\text{case.ids}}$ | | |
| ICC | | | | | | | 0.00 | | | | | | 0.00 | | | | | |
| N | $9164_{\text{case.ids}}$ | | | $9164_{\text{case.ids}}$ | | | $9164_{\text{case.ids}}$ | | | $9164_{\text{case.ids}}$ | | | $9164_{\text{case.ids}}$ | | | $9164_{\text{case.ids}}$ | | |
| Observations | 67030 | | | 67638 | | | 67638 | | | 67638 | | | 67638 | | | 67638 | | |
| Marginal $R^2$ / Conditional $R^2$ | NA | | | NA | | | 0.915 / 0.915 | | | NA | | | 0.899 / 0.899 | | | NA | | |

**Table S7. Half-lives of antibody titers for a given infection history, 2004-2023.**

| Infection History | Average titer magnitude at 1 year post last infection* | Slope(s)** | 95% Confidence Intervals | Half-lives *** | Inflection Point(s)**** |
|---|---|---|---|---|---|
| DENV Immune | 2.48[2.44,2.52] | **-0.12**<br>0.04 | **(-0.12, -0.11)**<br>(-0.002, 0.07) | 5.78<br>-17.33 | 8.79 |
| DENV | 1.87[1.83,1.92] | 0.02<br>**-0.05** | (-0.04, 0.08)<br>**(-0.06, -0.03)** | -34.66<br>13.86 | 2.3 |
| ZIKV | 0.85[0.81,0.89] | **0.30**<br>**-0.19** | **(0.28, 0.33)**<br>**(-0.23, -0.16)** | -2.31<br>3.65 | 3.2 |
| DENV-DENV | 2.71[2.61,2.80] | **-0.40**<br>**-0.04** | **(-0.55, -0.26)**<br>**(-0.16, -0.02)** | 1.73<br>17.33 | 2.31 |
| DENV-ZIKV | 3.01[2.98,3.13] | **-0.08**<br>-0.18 | **(-0.11, -0.05)**<br>(-0.89, 0.53) | 8.66<br>3.85 | 5 |
| ZIKV-DENV | 3.34[3.25,3.44] | **-0.28** | **(-0.35, -0.21)** | 2.48 | None |
| 2+DENV | 2.96[2.90,3.01] | **-0.36**<br>**-0.03** | **(-0.44, -0.29)**<br>**(-0.05, -0.02)** | 1.93<br>23.10 | 2.3 |
| 2+DENV-ZIKV | 3.56[3.44,3.67] | **-0.41**<br>0.16<br>-0.23 | **(-0.58, -0.23)**<br>(-0.05, 0.37)<br>(-0.49, 0.03) | 1.69<br>-4.33<br>3.01 | 2.4<br>4.4 |

*Predicted log10(DENV iELISA titer) and confidence intervals at 1 year after last infection.
** log10(DENV iELISA titer)/year. Significant slopes are bolded.
***Half-lives were calculated by computing log(2) divided by slope (log(2)/slope). Negative half-lives indicate an increase in titer magnitude.
**** Inflection points were calculated in years
